# GenBrain: A Generative Foundation Model of Multimodal Brain Imaging

**DOI:** 10.64898/2025.12.19.25342614

**Authors:** Chang Yang, Jianfeng Feng, Christian F. Beckmann, Stephen M. Smith, Weikang Gong

## Abstract

Neuroimaging faces a reproducibility crisis, where studies on small, heterogeneous datasets produce unreliable brain-wide associations and AI models that fail to generalize. To address this, we introduce GenBrain, a generative foundation model pretrained on approximately 1.2 million 3D scans from over 44,000 individuals across 34 imaging modalities to learn a population prior of brain structure and function. Crucially, GenBrain enables rapid, data-efficient adaptation, allowing any targeted study to generate biologically valid synthetic cohorts, conditioned on demographics, disease status, or other modalities, to augment statistical power and enhance generalizability. We demonstrate GenBrain’s transformative utility across 81 independent datasets spanning diverse populations, protocols, and clinical conditions. For image-level tasks, it achieves state-of-the-art performance in image enhancement and cross-modality synthesis while preserving subject-specific neurobiology. In population neuroscience, synthetic cohorts from GenBrain stabilize effect-size estimates and significantly improve the reproducibility of brain-wide association studies. For clinical AI, disease-specific fine-tuning of GenBrain substantially boosts the cross-site generalizability of prediction models. Finally, we prove its direct translational value when adapted to unseen modality and scarce clinical stroke data. GenBrain significantly improves predictions of acute stroke severity and chronic aphasia, demonstrating actionable utility under extreme data scarcity. By empowering small-scale studies with large-scale population priors, GenBrain provides a unified framework for more reproducible and clinically generalizable neuroimaging analysis.

## Introduction

Brain magnetic resonance imaging (MRI) has been instrumental in quantifying brain structure and functional organization^1^, as well as mapping neurodevelopmental and neurodegenerative trajectories in both health and disease^2^. A major goal is to reliably link inter-individual neuroanatomical and functional variations to clinical phenotypes, such as diagnostic status or symptom severity. However, the reproducibility of brain-wide association studies (BWAS) and the generalizability of AI models for disease diagnosis and phenotype prediction are severely hampered by the limited scale of most clinical studies^3-6^. This data scarcity stems from formidable practical constraints, including high acquisition costs, stringent governance restrictions, and patient privacy concerns. Consequently, research on rare diseases, specific patient populations, and multi-site clinical applications often relies on underpowered datasets, leading to unreliable biomarkers and models that fail to generalize to independent datasets^3,7,8^.

While large-scale brain imaging datasets, such as UK Biobank, have advanced population neuroscience^1,9^, they are not designed for specific diseases. Clinical translation requires neuroimaging methods that can amplify the value of smaller, targeted studies, which are often the only viable source of data for many clinical conditions. These studies face two major challenges. Firstly, at the imaging level, researchers often need to denoise and correct for artefacts in the acquired raw images to enhance their quality^10^. Additionally, they may need to impute missing modalities when certain scans of interest were either not included in the original study design or were difficult to acquire^11^. They may be also interested in studying modality relationships, such as brain structure vs. function^12^ and resting-state vs. task^13^. More importantly, for analysis tasks, studies generally aim to use brain images to examine associations between brain structure/function and cognitive or mental health phenotypes, or to predict non-imaging variables of interest, such as disease status. Current computational approaches address these problems in isolation. For instance, specialized models have been developed for image enhancement^10^, cross-modality synthesis^11,14^, or adapting predictive models from large to small datasets^15^.

We propose that a unified conditional generative model, pretrained on large-scale, high-quality brain imaging data, can address these challenges by encoding a strong prior of brain structure and function. Specifically, such a model would learn the multivariate distribution of imaging features across modalities as they appear in a general population. Once trained, the model would not only produce anatomically realistic images but, more critically, preserve subject-specific biological or pathological information when conditioned on imaging or non imaging phonetype of interests.

The generative model could be fine-tuned for image-level tasks like denoising scans, imputing missing sequences, by levegaging imaging as a condition. More importantly, it could synthesize imaging data conditioned on non-imaging variables, such as disease labels, by learning from small, targeted datasets while guided by the embedded prior. In this way, the model can efficiently infer how disease or lesions alter brain morphology and function based on its prior knowledge of general population distributions. Building on this capability, the framework would enable the generation of large, balanced synthetic cohorts for reproducible BWAS, as well as the augmentation of limited datasets to improve the robustness and generalizability of machine learning models in clinical neuroscience.

While a few recent studies have explored generative models for brain imaging, their scope has been limited to specific modalities (e.g., T1-weighted MRI^8,16^) or specific generation conditions (e.g., with text description^17^). Their validity is accessed on imaging-level tasks mainly, and often assessed only by low-level image quality metrics (such as PSNR and SSIM), with insufficient rigorous evaluation of their ability to preserve critical clinical biomarkers or to enhance downstream healthcare-relevant tasks. A comprehensive, multimodal generative framework, rigorously validated for its scientific and clinical utility, remains lacking.

In this study, we introduce GenBrain, a generative foundation model trained on approximately 1.2 million high-quality 3D brain images from the UK Biobank, spanning more than 44,000 subjects and 34 structural and functional modalities (Fig. 1). Built on a diffusion transformer framework, GenBrain overcomes key limitations of traditional GAN-based approaches, such as training instability and difficulty generating images of diverse dimensionalities^17,18^. GenBrain can synthesize high-quality structural (e.g., T1w, T2-FLAIR, susceptibility weighted imaging), diffusion (e.g., diffusion tensor imaging or neurite orientation dispersion and density imaging spatial maps), and functional (e.g., functional connectomes^19^, task activation maps) images or their derived voxel-wise feature maps, conditioned on demographic variables (e.g., age and sex) (Fig. 1a). Critically, these synthetic images are novel, do not replicate training data, and accurately reflect the biological information embedded in the conditioning variables (Fig. 1a). By serving as a strong population imaging prior, GenBrain can rapidly produce realistic synthetic images that adapt to limited data from different scanners and preprocessing pipelines, whether conditioned on demographics, phenotypes, disease status, or other imaging modalities (Fig. 1b). This enables it to: (1) Excel in image-level tasks (image enhancement, cross-modality synthesis) while uniquely preserving subject-specific biological signatures like age and sex effects; (2) Mitigate reproducibility challenges in BWAS by generating novel, biologically valid imaging and paired non-imaging data, thereby expanding the effective degrees of freedom for analysis; (3) Significantly improve cross-site generalizability of diagnostic classifiers through targeted synthetic data augmentation; (4) Extend to clinical applications, enhancing the prediction of acute stroke severity and chronic aphasia from held-out clinical-grade datasets, even under extreme data scarcity and novel imaging modalities (Fig. 1c).

**Fig. 1.**
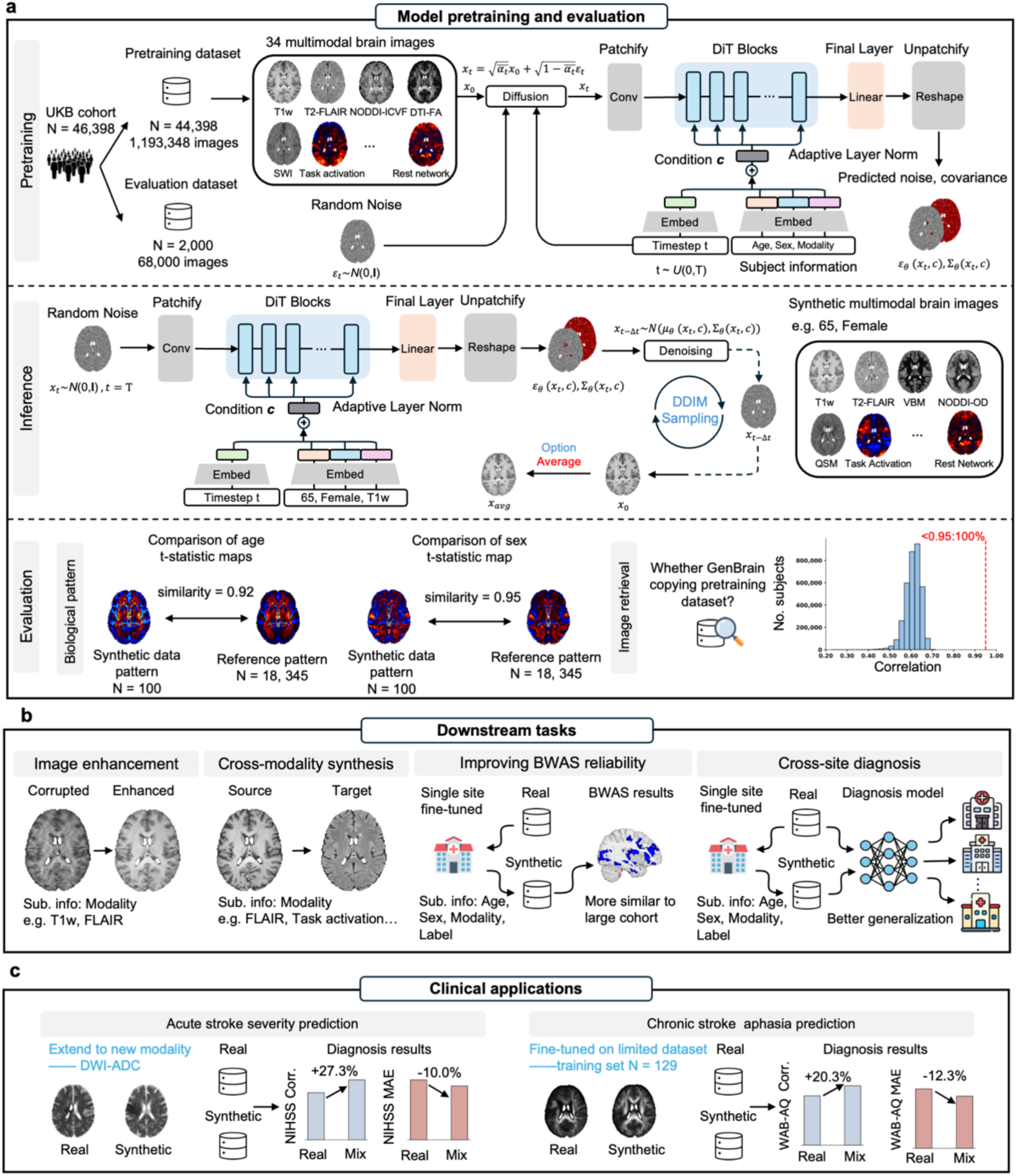
Overview of GenBrain foundation model. **a**, Model pretraining and evaluation. GenBrain was pretrained as a denoising diffusion probabilistic model on ∼1.2 million 3D brain images spanning 34 modalities from the UK Biobank (N=44,398 subjects). For inference, images are synthesized using denoising diffusion implicit model sampling, and generation fidelity can be enhanced by averaging multiple samples conditioned on the same input. Evaluations assess the preservation of biological patterns (e.g., age, sex) in synthetic images and the model’s ability to generalize beyond mere replication of the training set. **b**, Adaptation for downstream analysis tasks. The pretrained GenBrain model can be fine-tuned to enhance a spectrum of neuroimaging tasks, including image enhancement, cross-modality synthesis, improving the reliability of brain-wide association studies (BWAS), and augmenting cross-site generalizability in diagnostic machine learning. **c**, Translation to clinical applications. GenBrain robustly adapts to clinical-grade data, even with unseen modalities or extreme data scarcity. Synthetic data augmentation with GenBrain supports specific clinical predictions, such as acute stroke severity and chronic-phase aphasia.

By providing a single pretained model that enhances data quality, quantity, and analytical robustness, GenBrain establishes a foundation for more efficient, scalable, and clinically generalizable neuroimaging research.

## Results

### GenBrain generates morphologically realistic brain images

We began by assessing whether GenBrain could generate high-fidelity, multimodal brain images that capture fundamental biological variations. As we pretrain the model conditioning on age and sex, which are among the most significant phenotypic variables influencing brain morphology and function, we first tested whether these patterns were preserved in the generated multimodal images.

We first generated a population-level pseudo ground truth by performing a brain-wide association study on a large-scale reference cohort from the UK Biobank (N = 18,345) to establish voxel-wise t-statistic maps for age and sex effects (Methods). We then generated brain images for each modality, conditioned on the same age and sex parameters as a randomly selected subset (N = 100) of real images from the evaluation cohort. The cosine similarities between the t-statistic maps derived from the synthetic image maps and the reference maps were comparable to the similarities between the real image maps and the reference (mean relative age/sex similarity score: 88.2 ± 15.3% and 77.5 ± 21.1%) (Fig. 2a).

**Fig. 2.**
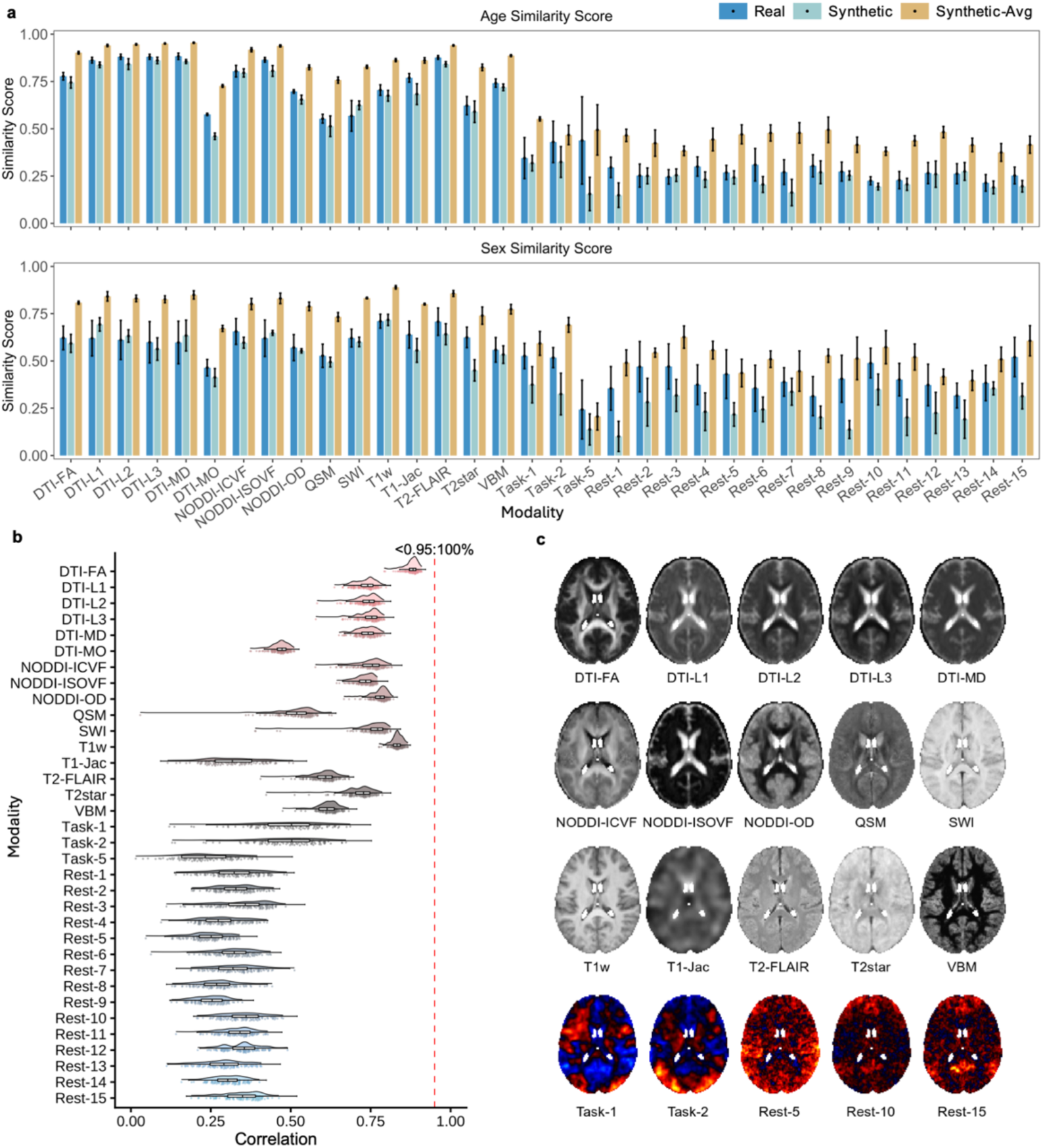
Evaluation of the pretrained GenBrain foundation model. **a**, Preservation of biological patterns in synthetic images. Age- and sex-related patterns were evaluated on randomly sampled small cohorts (N=100) of real data, synthetic data, and synthetic data averaged from five samples. The similarity score, defined as the cosine similarity between each small cohort’s voxel-wise t-statistic map and that of a large reference cohort (N=18,345), quantifies the preservation of biological patterns (higher scores indicate better preservation). Small cohorts were sampled five times with no overlap; error bars represent 95% confidence intervals. **b**, Image-retrieval analysis to assess memorization. Synthetic images were generated for all 34 modalities across 79 age–sex combinations represented in the pretraining data. For each synthetic image, we computed its Pearson correlation with every image of the same modality in the pretraining set. All pairwise correlations fell below a near-duplicate threshold (r = 0.95), demonstrating that the model generates novel images rather than replicating training data. For visualization, 100 correlations per modality were randomly sampled for the scatter plot. **c**. Example synthetic images for a age and sex label across multiple modalities (female, 65 years old).

To determine whether GenBrain could be rapidly adapted to learn other conditions of interest, we performed a similar experiment using fluid intelligence as a conditioning variable to fine-tune the mdoel (Supplementary Note 1). Results demonstrated that the model can also generate biologically reliable images when conditioned on variables with weaker overall brain association (Supplementary Fig. 1). These findings confirm that GenBrain captures complex, multivariate relationships between phenotypic variables and brain structure, rather than producing only anatomically plausible but biologically uninformative images.

### Averaging multiple samples enhances biological patterns within generated images

The signal-to-noise ratio of synthetic images could be improved by leveraging the precise spatial alignment afforded by nonlinear registration. By sampling multiple times from the diffusion model under identical conditions and computing their voxel-wise average, stochastic noise inherent in a single sample should average out, thereby enhancing the underlying biological signal. To test this, we generated five samples for each condition and averaged them to create a composite image. Re-evaluation of these averaged images against the above reference maps revealed a significant increase in cosine similarity for both age and sex effects (mean relative age/sex similarity score: 140.1 ± 26.6% and 128.7 ± 14.8%) (Fig. 2a), confirming that this averaging strategy substantially improves the functional quality and biological validity of the generated outputs, even surpassing the real images under the same sample size. The subsequent image-level analysis therefore used the averaged images.

### Synthetic images are novel and not replicas of training data

A critical consideration for generative models is the behaviour to memorize and reproduce individual samples from the training data. This memorization poses a serious risk to patient privacy and data compliance, and it undermines scientific utility by replicating idiosyncratic individual information rather than learning generalizable population statistics^20^.

To rigorously evaluate this, we conducted a comprehensive image-retrieval analysis. For each synthetic image, we computed the Pearson correlation coefficient with every image in the pre-training dataset across all 34 modalities and 79 age–sex combinations. We defined a stringent privacy-risk threshold at r > 0.95, indicating a near-identical copy. Not a single synthetic image exceeded this threshold (Fig. 2b). This demonstrates that GenBrain synthesizes novel image compositions that learn the underlying data distribution without simply memorizing or replicating individual training examples.

### GenBrain effectively enhances brain images while preserving biologically meaningful information

To evaluate GenBrain’s adaptability in downstram image-level tasks, we first repurposed its generative capabilities for the task of image enhancement. We fine-tuned the pre-trained GenBrain model (creating GenBrain-ft) on a limited dataset of 1,000 artificially corrupted T1w and 1,000 corrupted FLAIR images from the UK Biobank, paired with their original high-quality counterparts (see Methods). In this task, the model input was solely the low-quality image, without age or sex conditioning.

We compared GenBrain-ft against multiple baselines in the UKB evaluation set, including a population mean-brain method, an ablated version of GenBrain trained from scratch (GenBrain-st), the current state-of-the-art image enhancement method BME-X^10^, the general-purpose image-to-image translation model Pix2Pix^21^, and the T1w synthetic super-resolution method SynthSR^22-24^. GenBrain-ft demonstrated substantial improvements in standard image quality metrics (Fig. 3). For T1w images, it increased the peak signal-to-noise ratio (PSNR) from 23.75 dB to 26.45 dB (p = 2.03×10⁻^77^) and the structural similarity index (3D-SSIM) from 0.920 to 0.962 (p = 6.27×10⁻^45^) (Fig. 3a). For FLAIR images, PSNR increased from 20.45 dB to 22.23 dB (p = 1.00×10⁻^45^) and 3D-SSIM from 0.872 to 0.919 (p = 7.67×10⁻^78^), outperforming all other methods (Fig. 3b).

**Fig. 3.**
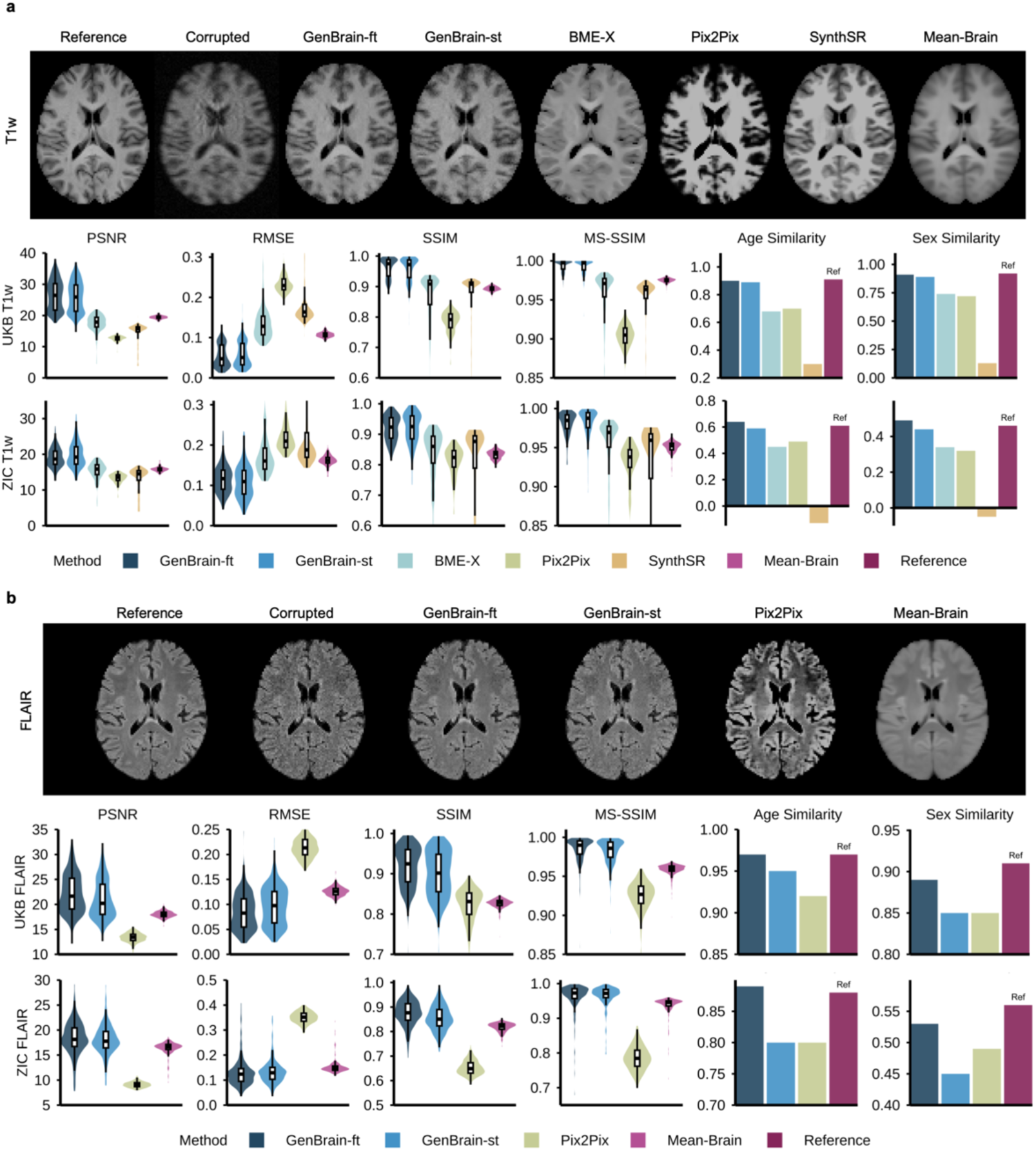
Fine-tuning GenBrain for image enhancement. **a**, T1-weighted image enhancement. The first row shows representative T1w images enhanced by different methods. The second and third rows report quantitative performance on the internal UK Biobank (UKB) evaluation set (N=500) and the external ZIC dataset (N=200), respectively. Performance is evaluated using image-level metrics (PSNR, RMSE, SSIM, MS-SSIM) and biological semantics-level metrics (Age/Sex Similarity), defined as the cosine similarity of voxel-wise t-statistic maps between the test cohort and a large reference cohort. **b**, T2-FLAIR image enhancement. The evaluation framework from **a** was applied to T2-FLAIR images, showing representative enhanced images, internal (UKB), and external (ZIC) performance.

Critically, beyond these traditional metrics, GenBrain-ft still excelled at restoring biologically relevant semantic information. The enhanced images recovered the age- and sex-related spatial patterns inherent in the original high-quality scans with high fidelity. GenBrain-ft significantly boosted age- and sex-similarity scores for T1w images from 0.76 to 0.90 (p = 0) and from 0.74 to 0.91 (p = 0) (Fig. 3a), and for FLAIR from 0.87 to 0.97 (p = 0) and from 0.79 to 0.89 (p = 0), consistently outperforming all other methods (Fig. 3b).

We further validated generalization on an external test set using the ZIC dataset (Fig. 3). Both GenBrain-ft and GenBrain-st achieved the best performance on image-level metrics (e.g., GenBrain-ft improved FLAIR PSNR from 16.43 dB to 18.38 dB, p = 1.76×10⁻^5^). However, GenBrain-ft maintained a distinct advantage in semantic fidelity, achieving an FLAIR age-similarity of 0.89 versus 0.80 (p = 0) and sex-similarity of 0.53 versus 0.45 (p = 0) compared with GenBrain-st (Fig. 3b). The above results demonstrated that pre-training is essential for robust generalization of semantic features across diverse datasets. The generate structural MRI images can be further super-resolved to higher resolution by applying a super resolution model GenBrain-SR (Supplementary Fig.3).

### GenBrain enables high-fidelity cross-modality synthesis with preserved biological signatures

We next evaluated whether GenBrain’s generative framework could be adapted for cross-modality synthesis, a critical task for imputing missing MRI sequences and enhancing dataset utility. We fine-tuned the pre-trained GenBrain model (GenBrain-ft) to map between modalities, including T1w-FLAIR synthesis, resting-state functional connectivity to task activation prediction, and structure-to-function mappings.

#### T1w-FLAIR synthesis

GenBrain-ft significantly outperformed models trained from scratch (GenBrain-st) and state-of-the-art specialized methods like TUMSyn^25^ and SynthSR across all quantitative metrics in the UKB dataset (Fig.4a). For T1w-to-FLAIR synthesis, GenBrain-ft generated structurally coherent FLAIR images, achieving a PSNR of 21.50 dB and a 3D-SSIM of 0.941. This surpassed the performance of all other methods, including GenBrain-st (PSNR = 20.38 dB, p = 4.57×10⁻^221^; 3D-SSIM = 0.928, p = 0), TUMSyn (PSNR =15.45 dB, p = 0; 3D-SSIM = 0.880, p=0), and U-Net (PSNR =17.41 dB, p = 0; 3D-SSIM = 0.916, p = 0). Most notably, the synthesized images preserved subject-specific biological information, with age-and sex-similarity scores (0.96 and 0.89) nearly matching those of the ground-truth scans (0.97 and 0.91). More importantly, we found that GenBrain-ft-synthesized T2-FLAIR images effectively retained white matter hyperintensities (WMH) features extracted from T1-weighted scans (Supplementary Fig. 2). In FLAIR-to-T1w synthesis task, GenBrain-ft again delivered the highest image fidelity (PSNR = 23.57 dB, 3D-SSIM = 0.958), outperforming GenBrain-st (PSNR = 23.04 dB, p = 2.55×10⁻^224^; 3D-SSIM = 0.950, p=0), TUMSyn (PSNR = 17.77 dB, p = 0; 3D-SSIM = 0.932, p = 1.70×10⁻^270^), SynthSR (PSNR = 16.21 dB, p = 0; 3D-SSIM = 0.911, p = 0), Pix2Pix (PSNR = 12.53 dB, p = 0; 3D-SSIM = 0.784, p = 0) and U-Net (PSNR = 22.12 dB, p = 8.29×10⁻^319^; 3D-SSIM = 0.945, p = 0). Its advantage in biological semantic fidelity was even more pronounced: it achieved age- and sex-similarity scores of 0.90 and 0.88, far exceeding the TUMSyn (0.41, 0.36, p = 0 and p = 0) and SynthSR (0.40, 0.17, p = 0 and p = 0), which failed to capture these critical ptterns (Fig. 4b).

**Fig. 4.**
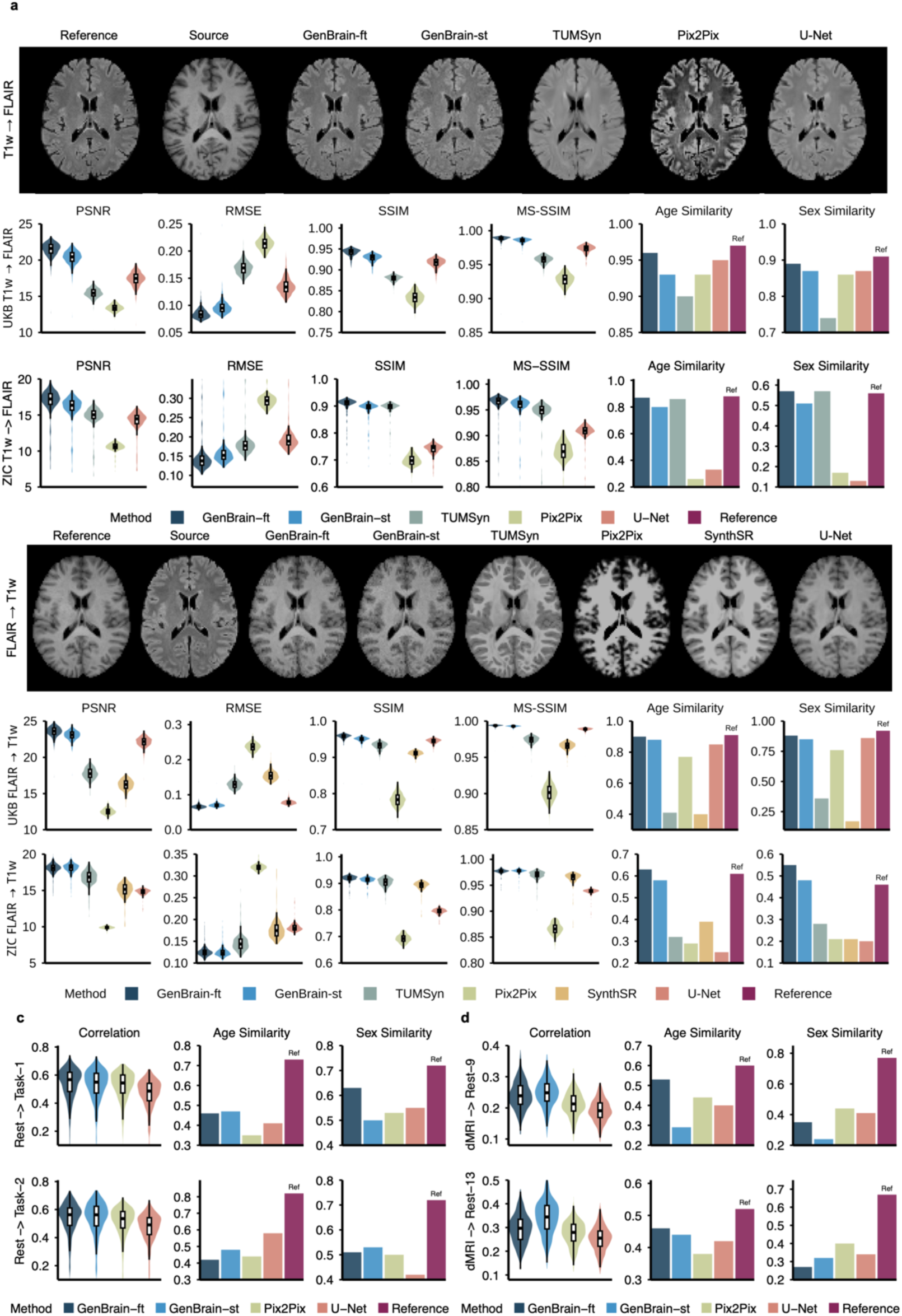
Fine-tuning GenBrain for cross-modality synthesis. **a,** T1w to T2-FLAIR synthesis. The first row shows representative synthetic T2-FLAIR images generated from T1w inputs by different methods. The second and third rows report quantitative synthesis performance on the internal UK Biobank (UKB) evaluation set (N=500) and the external ZIC dataset (N=200), respectively. Performance is evaluated using image-level metrics (PSNR, RMSE, SSIM, MS-SSIM) and biological semantics-level metrics (Age/Sex Similarity), defined as the cosine similarity of voxel-wise t-statistic maps between the test and a reference cohort. **b,** T2-FLAIR to T1w synthesis. The same evaluation framework from a was applied here, showing representative synthetic T1w images and their corresponding internal (UKB) and external (ZIC) performance. **c,** Resting-state to task-fMRI synthesis. Performance for synthesizing task-fMRI activation maps from resting-state functional connectivity patterns in UKB. The first and second rows report performance for the ‘shapes’ and ‘faces’ z-statistic maps, respectively, using spatial Pearson correlation and biological semantics-level metrics (Age/Sex Similarity). **d,** Diffusion MRI to functional connectivity synthesis. Performance for synthesizing seed-based functional connectivity maps (Language Network, Rest-9; Visual Central Network, Rest-13) from 9 diffusion MRI scalar maps (DTI: FA, MD, MO, L1, L2, L3; NODDI: OD, ICVF, ISOVF). Evaluations used the same metrics as in **c**.

External evaluation on the ZIC dataset confirmed the robustness of our approach. While all models experienced an expected performance drop on this independent cohort, GenBrain-ft maintained its relative superiority. It successfully synthesized high-quality images and was the only method to meaningfully preserve subject-specific age and sex patterns. For instance, in FLAIR-to-T1w synthesis, GenBrain-ft achieved age- and sex-similarity scores of 0.63 and 0.55, significantly higher than TUMSyn (0.32, 0.28, p = 0 and 0) and SynthSR (0.39, 0.21, p = 0 and 0) (Fig. 4b).

#### Resting-to-task prediction

We further challenged GenBrain with a complex resting-to-task mapping, i.e., predicting fMRI activation maps from resting-state functional connectivity patterns. Specifically, we fine-tuned GenBrain (GenBrain-ft) to translate seed-based functional connectivity maps (derived from DU15NET^19^, containing 15 cerebral cortical seeds across distinct functional networks) into z-statistic maps for two contrasts (shapes and faces) from an emotion task in the UK Biobank.

GenBrain-ft demonstrated superior performance in predicting subject-specific task activation patterns. It achieved the highest mean spatial correlation between the predicted and ground-truth maps (r = 0.54 for both contrasts), outperforming GenBrain-st on shapes contrast (r = 0.53, p = 2.01×10⁻^5^) and other established deep learning architectures on both contrast like Pix2Pix (r = 0.52/0.51, p = 3.80×10⁻^17^ and 2.99×10⁻^33^) and U-Net (r = 0.47/0.47, p = 4.85×10⁻^134^ and 1.37×10⁻^135^) (Fig. 4c). Crucially, the predictions generated by GenBrain-ft were not only accurate in their spatial patterns but also preserved meaningful biological information. The predicted task maps captured a significant portion of the age- and sex-related variance present in the actual task data. While the similarity scores for all methods were below the ground truth, GenBrain-ft still delivered competitive performance against other baseline methods (e.g., age/sex: GenBrain-ft: 0.46/0.63, Pix2Pix: 0.35/0.52 for shapes contrast, p = 0 and 0)(Fig. 4c).

#### Structure to function coupling

Finally, we evaluated whether GenBrain can effectively predicting patterns of functional connectivity from the brain’s white matter microstructure. We fine-tuned GenBrain to translate a multi-parametric set of nine diffusion MRI maps (including DTI and NODDI metrics) into seed-based resting-state functional connectivity maps. Performance was evaluated on predicting two distinct brain networks: the Language Network (Rest-9) and the Visual Central Network (Rest-13). The fine-tuned model (GenBrain-ft) demonstrated the most biologically plausible predictions of all models tested. In terms of spatial accuracy, GenBrain-ft achieved mean correlation scores of 0.24 for the Language Network and 0.29 for the Visual Central Network, performance on par with or exceeding the baseline models (e.g., Pix2Pix: 0.21 for Language, p = 1.27×10⁻^79^ and 0.28 for Visual Centeral, p = 1.94×10⁻^30^) (Fig. 4d). In addition, GenBrain-ft better captured subject-specific traits, achieving the highest age-similarity (0.53 for Language; 0.46 for Visual Centeral), compared with the second best (Pix2Pix: 0.44 for Language, p = 0; GenBrain-st: 0.44 for Visual Centeral, p = 1.53×10⁻^187^). (Fig. 4d).

### GenBrain mitigates small-sample bias and improves BWAS reproducibility

To directly validate GenBrain’s ability to address the reproducibility crisis in population neuroscience, we tested its capacity to stabilize BWAS impacted by limited data. We applied GenBrain’s core capability for data-efficient fine-tuning. Using a small target dataset, we fine-tuned the model conditioned on disease label and other key confounders (e.g., age, sex). This enabled the generation of a large and balanced synthetic cohort, effectively augmenting statistical power and mitigating confounder bias.

We then tested the central hypothesis: a BWAS performed on this augmented synthetic cohort would yield a Cohen’s *d* effect-size map that better approximates the ground truth from a large-scale reference cohort than a BWAS on the original small sample alone. We quantified map similarity using spatial correlation, Dice similarity coefficients (Dice), and a top-20% Dice metric focused on the most extreme effect sizes.

#### Schizophrenia

We first applied this framework to a multi-site voxel-based morphometry (VBM) dataset for schizophrenia (N = 2,831 from 17 sites). When fine-tuned on a small site (SH_JZ2, N = 330), BWAS results from the GenBrain-synthesized data showed marked improvement over the original site data. The spatial correlation with the large reference cohort increased from 0.17 to 0.29 (p = 0). The overall Dice coefficient improved from 0.59 to 0.69 (p = 0). This improvement was driven by enhanced overlap in both positive (0.35 to 0.38, p = 1.01×10^-128^) and negative (0.65 to 0.77, p = 0) effect regions. The top-20% Dice metric, reflecting agreement on the most salient effects, rose substantially from 0.19 to 0.29 (p = 0) (positive: 0.05 to 0.08, p = 3.73×10^-77^; negative: 0.20 to 0.30, p = 0). Critically, the effect-size maps derived from synthetic data matched to those of reference cohort, with much fewer high effect voxels shown up like noises. It also revealed biologically plausible patterns that were absent in the analysis of the original limited sample. These included decreased grey matter volumes in the third ventricle and hippocampus (Fig.5a), findings consistently reported in the literature on schizophrenia.

**Fig. 5.**
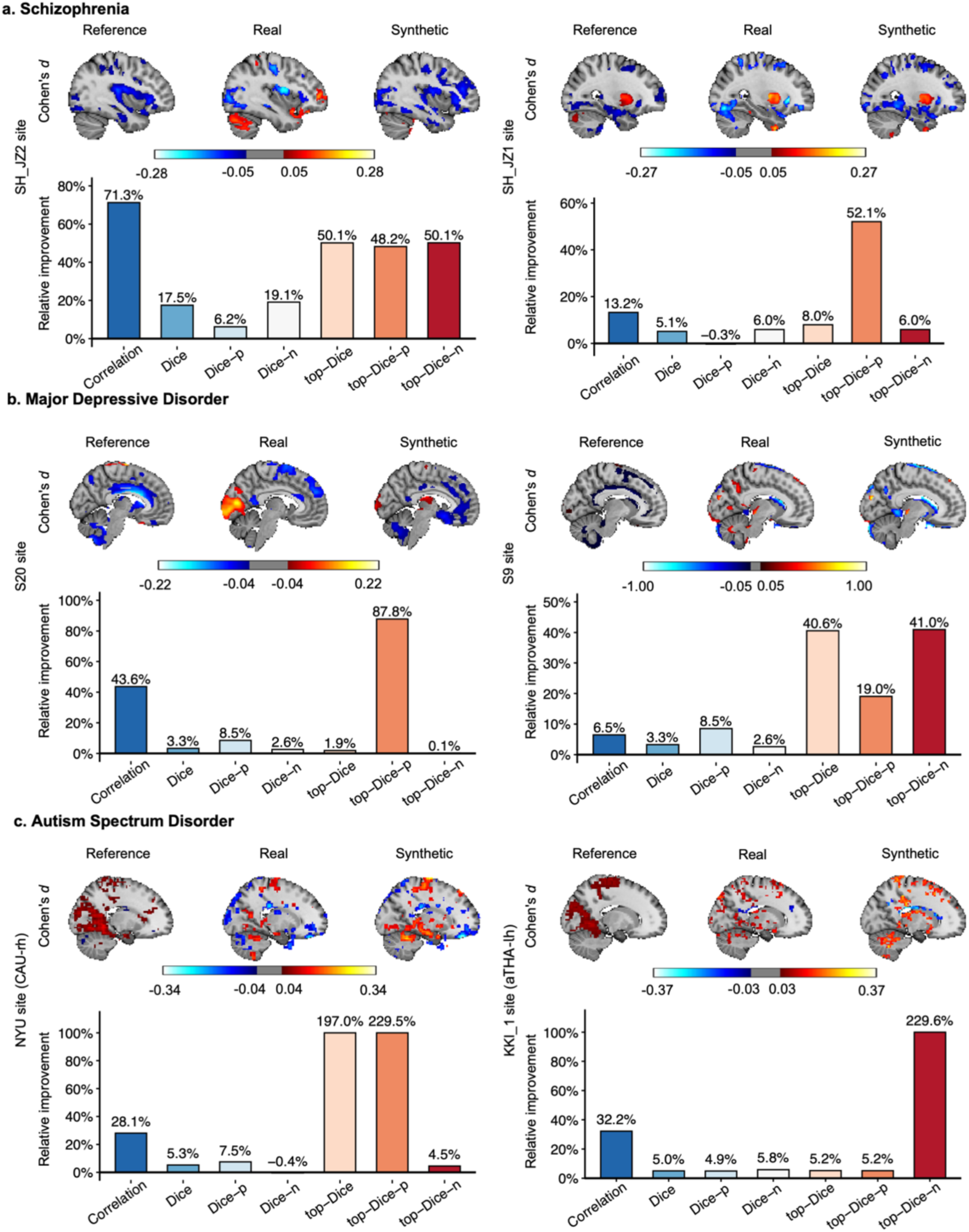
Enhancing brain-wide association study (BWAS) reliability with GenBrain. GenBrain was fine-tuned for case-control synthesis on three distinct brain disorders. For each disorder, results from two representative independent sites are shown. The model synthesizes case-control samples from a specific neuroimaging modality for BWAS to identify voxel-wise differences. Top Rows (**a-c**): For each site, the left panel shows the reference Cohen’s *d* map from a large multi-site cohort (excluding the fine-tuned site). The middle panel shows the Cohen’s *d* map from the real data of the fine-tuned site. The right panel shows the Cohen’s *d* map derived from the synthetic case-control samples generated for that site. Bottom Rows (**a-c**): Quantitative evaluation of how well the synthetic Cohen’s *d* maps aligns with the reference Cohen’s *d* maps compared to the real data. Bars indicate the relative improvement (percentage gain) of using synthetic over real data across seven metrics: Correlation: Pearson correlation between effect-size maps. Dice: Dice coefficient on the full Cohen’s *d* map (micro-averaged across positive and negative effects). Dice-p / Dice-n: Dice coefficient for positive (*d* > 0) / negative (*d* < 0) effects. Top-Dice (overall): Dice coefficient for the top 20% of |Cohen’s *d*| values (micro-averaged). Top-Dice-p / Top-Dice-n: Dice coefficient for the top 20% of positive / negative effects. **a,** Schizophrenia. Fine-tuned on VBM modality from the SH_JZ2 site (N=330) and SH_JZ1 site (N=298). **b,** Major Depressive Disorder. Fine-tuned on VBM modality from the S20 site (N=533) and S9 site (N=100). **c,** Autism Spectrum Disorder. Fine-tuned on subcortical seed-based functional connectivities (modalities unseen in pretraining) from the NYU site (N=171; right caudate nucleus seed) and KKI_1 site (N=198; left anterior thalamus seed).

The robustness of this approach was further validated by repeating the fine-tuning and analysis on other independent sites. On the SH_JZ1 site (N = 298), using GenBrain-synthesized data for analysis yielded consistent gains, with the spatial correlation increasing from 0.27 to 0.30 (p = 1.06×10^-220^), the overall Dice coefficient from 0.64 to 0.68 (p = 0), and the top-20% Dice metric from 0.25 to 0.27 (p =2.34×10^-262^). Similar improvements were observed on the zhengzhou site (N = 253), where correlation rose from 0.39 to 0.41 (p = 0), Dice coefficient improved from 0.63 to 0.64 (p = 1.25×10^-165^), and on the extreme small fBIRN site (N = 107), where correlation improved from 0.13 to 0.15 (p = 1.70×10^-104^).

#### Major Depressive Disorder (MDD)

In the analysis of VBM data from a multi-site MDD cohort (N =2,831 from 24 sites), GenBrain again demonstrated its utility. When fine-tuned on a site where the original effect-size map exhibited a moderate positive correlation with the reference (S20 site, N = 533), the synthetic data yielded a BWAS Cohen’s *d* map with improved alignment with the large reference cohort (Fig. 5b). The spatial correlation increased from 0.13 to 0.18 (p = 2.08×10^-131^), and the overall Dice coefficient improved from 0.65 to 0.68 (p = 1.16×10^-53^), with gains in both positive (0.34 to 0.36, p = 4.95×10^-48^) and negative (0.74 to 0.78, p = 6.06×10^-38^) effect regions. The top-20% Dice metric for the most salient positive effects remained same at 0.19, and positive regions improved from 0.03 to 0.06 (p = 8.40×10^-98^).

On the smaller S9 site (N=100), where the original effect-size map showed only a weak positive correlation (r = 0.01) with the reference, the GenBrain-based analysis achieved a comparable correlation but markedly improved spatial consistency (Fig. 5b): the overall Dice coefficient rose from 0.46 to 0.60 (p = 2.00×10^-219^), driven primarily by the negative effect regions (0.50 to 0.69) (p = 5.41×10^-223^), and the top-20% Dice increased from 0.11 to 0.16 (p = 1.59×10^-175^). Likewise, on the S1 site (N =148), correlation changed marginally (0.11 to 0.09), yet Dice improved from 0.54 to 0.57 (p = 5.73×10^-15^).

Conversely, on the S8 site (N = 150), where the original effect-size map exhibited a weak negative correlation with the reference (r = –0.04), using the site’s own data for fine-tuning proved ineffective (r = –0.06). This result underscores a key principle: the success of fine-tuning approach is contingent upon the initial site data capturing at least a minimal, population-consistent pattern of effects. This finding suggests that for sites with highly aberrant or weak signals, strategies such as multi-site fine-tuning or quality screening prior to synthesis are necessary to ensure robust performance.

#### Autism Spectrum Disorder (ASD)

To further assess the generalizability of the framework across fundamentally different imaging modalities, we applied it to functional connectivity (FC) maps derived from the combined ABIDE I and II datasets. Sixteen seed-based FC maps were generated across eight subcortical regions (left and right hemispheres) using the Tian subcortical atlas^26^, a set of new modalities entirely unseen during GenBrain’s pre-training, thus presenting a significant generalization challenge. The results closely mirrored our structural findings, indicating that GenBrain can effectively generalize to new functional-level representations.

On the NYU site (N = 171), GenBrain-synthesized data led to widespread improvements across multiple seed-based FC maps (Fig. 5c). For example, in the *putamen* (PUT-rh and PUT-lh; Tian indices 7 and 15) seed-based maps, spatial correlation increased from 0.12 to 0.18 (p = 6.87×10^-235^) for PUT-rh and from 0.12 to 0.15 (p = 3.74×10^-194^) for PUT-lh. The Dice coefficient improved from 0.51 to 0.56 (p = 4.46×10^-210^) and from 0.51 to 0.57 (p = 1.17×10^-175^), while the top-20% Dice metric rose substantially from 0.11 to 0.22 (p = 3.59×10^-251^) and from 0.12 to 0.22 (p = 5.20×10^-183^).

On the KKI_1 site (N = 198), GenBrain-synthesized data similarly produced widespread improvements across seed-based FC maps (Fig. 5c). For example, in the *anterior thalamus* (aTHA-rh and aTHA-lh; Tian indices 4 and 12), correlation increased from 0.11 to 0.14 (p = 3.95×10^-70^) and from 0.16 to 0.22 (p = 6.60×10^-186^), respectively. The Dice coefficient improved from 0.66 to 0.67 (p = 2.53×10^-72^) and from 0.68 to 0.71 (p = 4.81×10^-213^).

Across both datasets, the *caudate nucleus* (CAU-rh and CAU-lh; Tian indices 8 and 16) showed particularly robust enhancements. On the NYU site, correlations increased from 0.21 to 0.27 (p = 2.10×10^-246^) and from 0.14 to 0.21 (p = 5.57×10^-266^), with Dice coefficients improving from 0.57 to 0.60 (p = 2.00×10^-125^) and from 0.58 to 0.59 (p = 7.16×10^-190^), and the top-20% Dice metric markedly rising from 0.10 to 0.31 (p = 7.75×10^-203^) and from 0.14 to 0.24 (p = 1.43×10^-208^). On the KKI_1 site, correlations improved from 0.09 to 0.13 (p = 3.42×10^-179^) and from –0.02 to 0.09 (p = 0), with Dice increasing from 0.56 to 0.57 (p = 6.34×10^-143^) and from 0.51 to 0.54 (p = 6.68×10^-296^), and top-20% Dice from 0.16 to 0.17 (p = 1.63×10^-163^)and from 0.11 to 0.14 (p = 0).

Together, these results demonstrate that GenBrain substantially enhances the reproducibility and biological plausibility of functional connectivity analyses across independent sites and previously unseen functional modalities.

### GenBrain enhances cross-site generalizability of neuroimaging-based disease diagnosis

Beyond enhancing population level analysis, a principal challenge in deploying machine learning models for clinical neuroimaging is their limited generalization to data from unseen scanners and sites, a problem often exacerbated by small sample sizes and limited image diversities. To overcome this, we developed a strategy that leverages the generative model. We hypothesized that by fine-tuning GenBrain on a disease cohort (with disease status as an additional condition) and using it to synthesize individualized, high-quality, label-preserving images, we could effectively augment training datasets and significantly improve the cross-site robustness of diagnostic classifiers.

We evaluated this approach on two distinct neuropsychiatric disorders: schizophrenia and Alzheimer’s disease. For each, we trained site-specific classifiers on combinations of real and synthetic data and rigorously assessed their performance in within-site and cross-site validation paradigms.

#### Schizophrenia classification

We first addressed the binary classification of patients with schizophrenia versus healthy controls using T1-weighted data from a multi-site consortium. After fine-tuning GenBrain on data from a single training site (conditioned on age, sex, and disease status), we generated synthetic images to augment the original training set at synthetic-to-real ratios from 0:1 (real data only) to 3:1. Classifiers (LightGBM^27^) were trained on 32 regional volumetric features extracted from both real and synthetic images. We evaluated performance across 7 sites, comprising 42 unique cross-site pairs and 7 within-site pairs (Methods).

The inclusion of GenBrain-synthesized data markedly improved the model’s ability to generalize to external, unseen sites (Fig. 6a). Classifiers trained on augmented data demonstrated significantly higher mean cross-site accuracy compared to the baseline models trained on real data alone (mean accuracy increase: 7.34%, two-sided p = 2.85×10⁻^3^). Specifically, 26 of 42 (61.9%) cross-site predictions showed statistically significant improvement, while 5 (11.9%) pairs showed no significant change and 11 (26.2%) showed a slight decrease, indicating robust positive transferability. Data augmentation also benefitted within-site classification. Models trained and tested on data from the same site (via cross-validation) exhibited improved accuracy at 6 of the 7 sites upon the addition of synthetic data (e.g., fBIRN site: accuracy 0.59 to 0.68, p = 2.85×10⁻^80^; zhengzhou site: accuracy 0.76 to 0.80, p = 6.04×10⁻^100^).

**Fig. 6.**
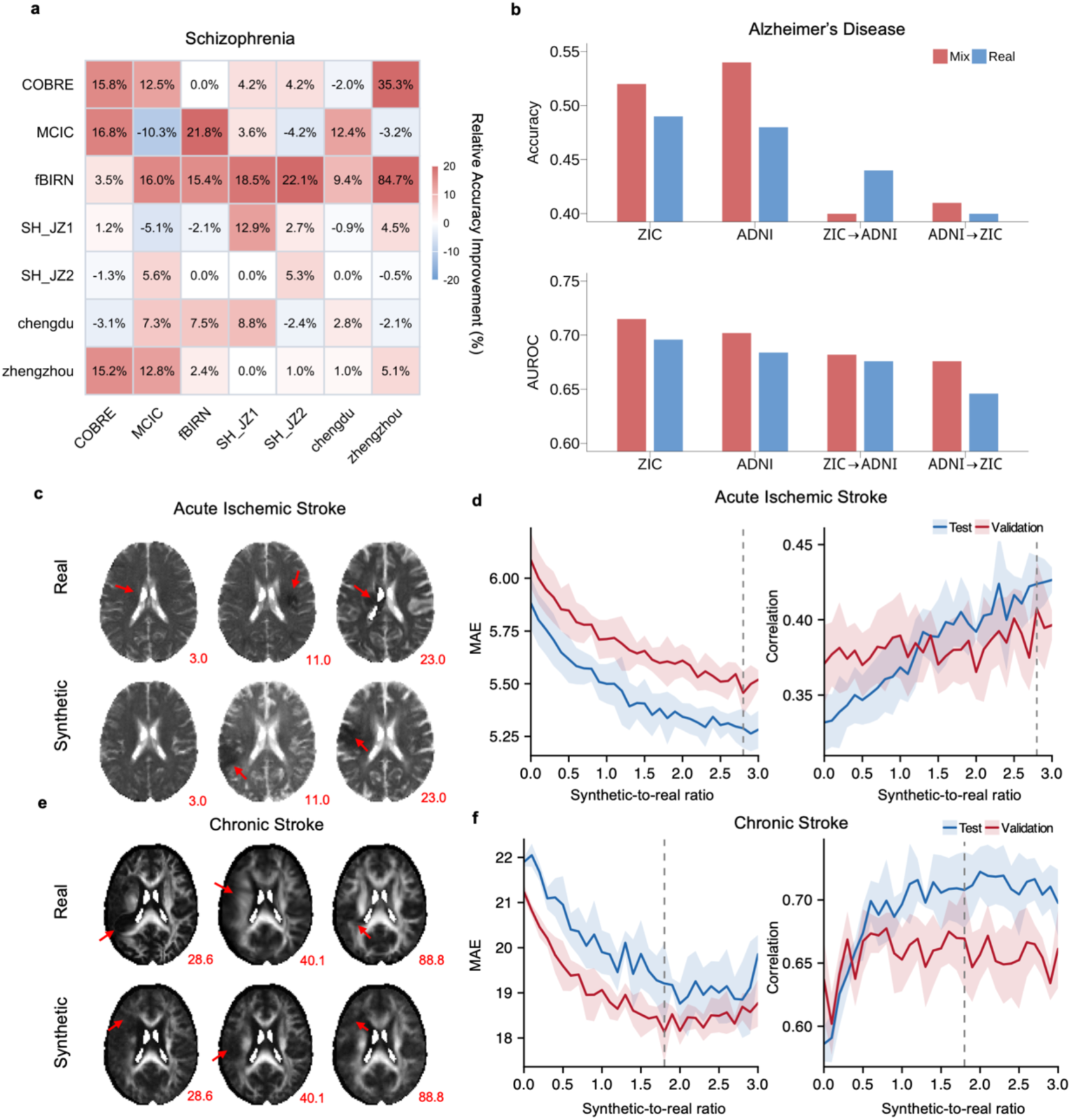
Synthetic-image data augmentation for cross-site diagnosis and clinical applications. **a**, Schizophrenia diagnosis. Cross-site binary classification performance across seven independent datasets. Each element shows the relative improvement in accuracy when augmenting the training data of source site with synthetic images generated by GenBrain versus using real data alone. Rows represent source site and columns represent target site. **b**, Clasification of Alzheimer’s disease, MCI and normal controls. Cross-site three-class classification performance on the ADNI and ZIC datasets. Bar graphs report the improvement in classification accuracy and AUROC achieved with synthetic data augmentation. **c**, Example acute ischemic stroke images. Representative real and synthetic DTI–ADC image pairs, matched for age, sex, and NIH Stroke Scale (NIHSS) score (reported below each image). **d**, Acute stroke severity prediction. Model performance in predicting NIHSS scores on validation and test sets. Curves show the Pearson correlation (left) and mean absolute error (MAE, right) as a function of the synthetic-to-real data ratio used during training. The optimal synthetic-to-real data ratio, determined by the minimum mean validation MAE, is indicated by a dashed vertical line. **e**, Example chronic stroke images. Representative real and synthetic DTI–FA image pairs, matched for age, sex, and Western Aphasia Battery–Aphasia Quotient (WAB-AQ) (reported below each image). **f**, Chronic stroke aphasia prediction. Model performance in predicting WAB-AQ scores on validation and test sets, showing Pearson correlation and MAE across varying synthetic-to-real data ratios.

The advantage of our method was most pronounced at sites with limited training data. For instance, at the COBRE site, which contained only 99 subjects in the training set, the baseline model trained directly on this small cohort exhibited poor cross-site generalization (e.g., achieving an accuracy of 0.46 on the zhengzhou site and 0.59 on the MCIC site). In contrast, augmenting the training set with GenBrain-synthesized images substantially improved performance across these sites (Fig.6a; accuracy 0.62 on zhengzhou, p = 0; accuracy 0.67 on MCIC, p = 0).

#### Alzheimer’s disease classification

We next evaluated our method on a three-class classification task to distinguish between patients with Alzheimer’s disease, mild cognitive impairment, and cognitively normal controls. Using the same strategy applied to schizophrenia, we fine-tuned GenBrain and augmented training sets with synthetic data before training LightGBM classifiers. Experiments were conducted using the ZIC (N = 1,136) and ADNI^28^ (N = 2,632) datasets to assess within- and cross-site performance.

The inclusion of GenBrain-synthesized data consistently enhanced classification performance on both datasets (Fig. 6b). The optimal synthetic-to-real data ratio was found to be dataset-dependent: On the ZIC dataset, peak performance occurred at a ratio of 0.8:1, increasing accuracy from 0.49 to 0.52 (p = 1.09×10^-135^) and area under the receiver operating characteristic curve (AUROC) from 0.696 to 0.715 (p = 2.16×10^-144^). On the ADNI dataset, the optimal ratio was 0.3:1, boosting accuracy from 0.48 to 0.54 (p = 0) and AUROC from 0.684 to 0.702 (p = 2.88×10^-258^). For cross-site enhancement, when trained on ADNI and tested on ZIC, the model showed a substantial improvement, with AUROC increasing from 0.646 to 0.676 (bootstrap p = 0). In the reverse direction, we observed a modest gain in AUROC from 0.676 to 0.682 (bootstrap p = 2.77×10^-203^), while other performance metrics remained stable. These results demonstrates that our augmentation method can enhance model robustness across heterogeneous sites without compromising baseline performance.

### Adapting GenBrain to clinical-grade datasets for enhanced outcome prediction

A critical test of a medical AI model’s utility is its robustness and generalizability across held-out datasets with varying acquisition protocols and patient pathologies. We rigorously evaluated GenBrain’s ability to augment machine learning in two challenging, clinically representative scenarios: prediction of acute stroke severity and chronic aphasia quotient, using entirely unseen clinical-grade datasets and modalities not encountered during pre-training.

#### Synthetic data augmentation improves acute stroke severity prediction

The National Institutes of Health Stroke Scale is a standardized tool for assessing neurological deficit, guiding acute treatment decisions, and estimating prognosis^29^. To test GenBrain’s utility in this acute setting, we utilized diffusion tensor imaging–apparent diffusion coefficient (DTI-ADC) maps from 1,106 participants in the Stroke Outcome Optimisation Project, a modality absent from GenBrain’s pre-training data. Note that DTI-ADC is also a modality that is never seen in the GenBrain’s pretraining.

A baseline 3D ResNet^30,31^ regression model trained exclusively on real DTI-ADC data achieved a test mean absolute error (MAE) of 5.88 and a prediction correlation of r = 0.33. We then systematically augmented the training set with GenBrain-synthesized DTI-ADC images, increasing the synthetic-to-real data ratio. The incorporation of synthetic data yielded consistent and significant performance improvements (Fig. 6c, d). The MAE decreased monotonically and correlation increased across all tested ratios. The optimal performance was observed at a synthetic-to-real ratio of 2.8:1, which corresponded to a test MAE of 5.29 (a 10.0% reduction, p = 9.25×10⁻⁵) and a prediction correlation of r = 0.42 (a 27.3% improvement, p = 2.42×10⁻^3^), significantly outperforming the baseline model.

#### Augmentation mitigates data scarcity in chronic aphasia prediction

We next evaluated GenBrain in a data-scarce scenario: predicting the Western Aphasia Battery–Aphasia Quotient, which is a comprehensive metric of language impairment in chronic stroke. We used fractional anisotropy (FA) maps from 233 participants in the Chronic Aphasia Recovery Cohort.

The baseline model, trained on a limited set of real FA scans (N=129), achieved a test MAE of 21.90 and a correlation of r = 0.59. Progressive augmentation with synthetic FA images markedly enhanced prediction accuracy (Fig. 6f). Performance gains were evident across most ratios, with the optimal validation result at a 1.8:1 synthetic-to-real ratio. This model achieved a test MAE of 19.21 (a 12.3% reduction, p = 3.46×10⁻^3^) and a correlation of r = 0.71 (a 20.3% improvement, p = 2.50×10⁻^3^) over the baseline. Performance plateaued at higher ratios, indicating a well-defined optimum for data augmentation.

Together, these results demonstrate that synthetic neuroimaging data generated by GenBrain can meaningfully augment distinct, clinical-grade datasets. The framework significantly boosts the accuracy of clinical outcome predictions for both acute and chronic stroke, even when applied to previously unseen modalities and under conditions of inherent data scarcity. This ability to extrapolate to new clinical contexts and mitigate data limitations underscores GenBrain’s potential for scalability and broader applicability in medical AI.

## Discussion

We introduced GenBrain, a generative foundation model for multimodal neuroimaging, pretrained on approximately 1.2 million 3D scans across 34 structural and functional modalities. Our work was motivated by the central challenge of modern neuroimaging: a reproducibility crisis stemming from small, heterogeneous clinical datasets, which limit the discovery of robust brain-wide associations and the generalizability of predictive models. We posited that a single model, encoding a strong population prior of brain structure and function, could be rapidly adapted to overcome this fundamental limitation. As demonstrated, fine-tuning GenBrain on small, target-specific datasets enables the generation of high-fidelity synthetic cohorts that preserve crucial biological and pathological signatures. This capability directly enhances the quality and quantity of imaging data, leading to more reliable, reproducible, and powerful downstream analyses across population neuroscience and clinical AI applications.

A key finding is that pretraining on population-scale data enable GenBrain to learn a rich prior of brain morphology and function that generalizes robustly across domains. This was not merely a matter of improving traditional image quality metrics like PSNR and SSIM. More importantly, GenBrain-generated images consistently captured underlying biological variation, such as age,sex and disease effects, even when the model was not explicitly conditioned on these demographics during tasks like image enhancement. This semantic fidelity, the preservation of meaningful phenotypic information, is a critical advantage over task-specific models trained from scratch, which often fail to learn these complex, multivariate relationships from limited data. This capability was further enhanced by our multi-sample averaging strategy, which effectively amplified the biological signal while suppressing stochastic noise, a technique uniquely enabled by the precise spatial alignment of neuroimaging data.

The utility of this semantically-rich generation was proven across diverse applications. In image-level tasks, GenBrain excelled at image enhancement and cross-modality synthesis, outperforming specialized state-of-the-art models. Crucially, it was the method that the most successfully preserved subject-specific biological patterns (e.g., age/sex effects) in synthesized images, turning modality imputation from a purely structural task into a biologically meaningful one.

More importantly, in population neuroscience, GenBrain directly addresses the reproducibility crisis in BWAS, a domain historically plagued by underpowered designs^3-5^. Our method mitigates small-sample bias and augments statistical power. As experimentally validated, fine-tuning GenBrain on a small target dataset enabled the generation of a large, confounder-balanced synthetic cohort. A BWAS performed on this augmented cohort produced effect-size maps that were significantly more reproducible and better aligned with statistical parametric maps obtained from large-scale reference cohorts than analyses confined to the original small sample. This approach aligns with recent findings on the importance of study design for BWAS reproducibility. Rather than altering study design^4^, a practical challenge with constrained resources, GenBrain provides a direct solution by synthetically enabling well-powered, balanced designs.

For predictive modeling, we leveraged GenBrain’s capacity for rapid, data-efficient fine-tuning. By conditioning the model on clinical labels (e.g., schizophrenia, Alzheimer’s disease) alongside key confounders, we generated large, balanced synthetic cohorts for targeted data augmentation. This strategy substantially improved the cross-site generalizability of diagnostic classifiers. The benefit was most pronounced for sites with the smallest sample sizes, demonstrating GenBrain’s potential to overcome the critical data scarcity that hampers clinical AI. This direcly demonstrated GenBrain’s ability to generate biologically plausible "new" and "individualized" samples, drawn from the estimated distribution of true populations, ensures it moves beyond simply generating denoising images or modeling an average brain.

Furthermore, GenBrain also demonstrated remarkable extensibility, successfully generating previously unseen modalities (e.g., DTI-ADC, FA maps from clinical stroke cohorts) and enhancing the prediction of clinically relevant outcomes like stroke severity and chronic aphasia, even in data-scarce scenarios.

A major concern with any image generative model is its tendency to memorize training data, posing privacy risks. Our rigorous image-retrieval analysis confirmed that the pretrained GenBrain synthesizes novel images, learning the underlying population distribution without replicating individual training examples. This ensures patient privacy and validates the model for scientific use.

Despite its broad success, certain limitations highlight avenues for future work. First, while effective, the performance in complex tasks like predicting task-fMRI from resting-state data, though state-of-the-art, did not fully capture the ground-truth biological variance, indicating room for improvement in modeling high-level cognitive states. Second, the benefits of synthetic data augmentation exhibited diminishing returns beyond an optimal ratio, emphasizing that synthetic data are a powerful complement to, not a replacement for, real acquisitions. Third, performance gains were contingent on the quality of the fine-tuning data, i.e., sites with signals highly discordant from population priors saw limited improvement, suggesting the need for data quality screening or multi-site fine-tuning strategies.

In conclusion, GenBrain establishes that a foundation model approach, pretrained on population-scale data and flexibly fine-tuned, can serve as a versatile engine for enhancing neuroimaging research. It provides a unified solution to improve image quality, impute missing modalities, augment datasets for machine learning, and increase the statistical rigor of brain-wide association studies. By converting large-scale priors into actionable, task-specific synthetic data, GenBrain offers a practical and powerful path toward more data-efficient, reproducible, and generalizable brain imaging science.

## Data availability

The individual-level neuroimaging data used in this study were obtained from UK Biobank (https://www.ukbiobank.ac.uk/), Schizophrenia and ZIC-AD (https://zib.fudan.edu.cn/), MDD (https://rfmri.org/REST-meta-MDD), ADNI(https://adni.loni.usc.edu/), ABIDE (https://fcon_1000.projects.nitrc.org/indi/abide/), SOOP (https://openneuro.org/datasets/ds004889/versions/1.1.2), and ARC (https://openneuro.org/datasets/ds004884/versions/1.0.2).

## Code availability

The code to reproduce the results can be accessed at **GitHub**.

## Author contributions

C.Y, W.G designed the project, collected and analyzed the data, wrote the manuscript. C.Y, W.G, J. F, C.B, S.S discussed the results and revised the manuscript.

## Acknowledgements

We thank all participants for contribution of data. The UKB analyses were conducted using the UKB Resource under application no. 19542. The computations in this research were performed using the CFFF platform of Fudan University.

## Methods

### Datasets for pretraining and internal evaluation

We used the population-scale brain imaging resource from the UK Biobank (UKB). Our study included 46,398 participants, yielding a total of 1,261,348 3D brain magnetic resonance imaging (MRI) scans across 34 distinct imaging modalities. From this, a cohort of 44,398 participants (1,193,348 scans) was used for model pretraining. An independent hold-out set of 2,000 participants (68,000 scans) was reserved for internal evaluation.

UKB imaging data were preprocessed primarily based on FSL^32^ and FreeSurfer^33^ by using an optimized pipeline^34^. For this study, we utilized 3D voxel-wise images from all 34 modalities, which were registered to the MNI152 2mm standard space before being fed to GenBrain. The modalities comprised: (1) Resting-state functional MRI (fMRI): 15 seed-based functional connectivity maps based on DU15NET^19^(A novel 15-network parcellation of the cerebral cortex). (2) Task-based fMRI: 3 contrast maps from an emotion task (shapes, faces, faces > shapes). (3) Diffusion MRI: 9 maps, including 6 from diffusion tensor imaging (DTI; FA, MD, MO, L1, L2, L3) and 3 from neurite orientation dispersion and density imaging (NODDI; OD, ICVF, ISOVF). (4) Structural T1-weighted MRI: 3 maps, including the original T1w image, grey matter volume from voxel-based morphometry (VBM), and the Jacobian map (reflecting local volume change after nonlinear normalization). (5) Susceptibility-weighted MRI: 3 maps, including the original SWI, T2-star, and quantitative susceptibility mapping (QSM). (6) Structural T2-FLAIR images. A complete description of all modalities is provided in Supplementary Table 1. In addition to operating in the MNI152 2 mm standard space, GenBrain can be fine-tuned to MNI152 1 mm standard space to support tasks such as image super-resolution and cross-modality synthesis (Supplementary Note 4-5).

### Downstream evaluation datasets

To rigorously evaluate the generalizability of GenBrain, we conducted extensive downstream analyses on 81 external, publicly available datasets, totaling 12,645 subjects across five major neurological and psychiatric conditions. The use of these datasets, which were processed with heterogeneous pipelines, tests the model’s robustness to real-world variability. The datasets include: (1)Alzheimer’s Disease (AD): T1-weighted images from the Alzheimer’s Disease Neuroimaging Initiative (ADNI; N=2,632) and T1w/T2-FLAIR images from the ZIC dataset (N=1,136). (2) Schizophrenia (SCZ): T1w images and the derived Voxel-based morphometry (VBM) maps derived from T1w images across 17 independent sites (N=2,958) from the ZIB consortium^35^. (3) Major Depressive Disorder (MDD): VBM maps from 24 sites (N=2,831) obtained from the REST-meta-MDD consortium. (4) Autism Spectrum Disorder (ASD): Resting-state fMRI data from the combined ABIDE I and II cohorts^36,37^ (36 sites; N=1,778). From each scan, we derived 16 new seed-based functional connectivity maps from the Tian subcortical alats^26^, which were unseen during pretraining. (5) Stroke: Diffusion MRI maps from two cohorts: acute stroke cases from the SOOP dataset (DTI-ADC maps; N=1,106)^38^ and chronic stroke cases from the ARC dataset (FA maps; N=213)^39^. The preprocessing of external T1w data followed the official UKB structural pipeline for consistency. VBM data for the MDD analysis were processed using the SPM-based pipeline released by the REST-meta-MDD project^40^. ABIDE resting-state fMRI data were preprocessed by the Preprocessed Connectomes Project using DPABI^41^. Diffusion maps (DTI-ADC) were aligned to MNI152 2mm standard space using the SynthMorph tool in FreeSurfer. Note that in some of the external evaluation datasets we used different data preprocessing pipelines to test the robustness of GenBrain.

### Model Pretraining: Denosing diffusion probabilistic model

The denoising diffusion probabilistic model (DDPM)^42^ provides a generative framework in which clean data are gradually corrupted with Gaussian noise in the forward process, and a neural network is trained to reverse this trajectory by predicting and removing the noise at each timestep. Training minimizes the mean squared error between the predicted and true noise and the per-step KL divergence between the true posterior and the model reverse kernel, yielding high-fidelity samples, but DDPM typically requires hundreds to thousands of denoising iterations for generation. To improve efficiency, the denoising diffusion implicit model (DDIM)^43^ reformulates the reverse process as a deterministic trajectory, enabling high-quality generation with significantly fewer sampling steps.

Building on this framework, GenBrain employs DDPM training to learn the noise-prediction process directly from 3D brain voxels, while adopting DDIM-based sampling for efficient inference. During generation, the model samples in 50 steps and applies classifier-free guidance (scale = 1.2) to balance conditional fidelity and diversity, enabling the synthesis of high-quality brain images conditioned on demographic and modality information.

### Model Pretraining: Diffusion Transformer

The Diffusion Transformer (DiT)^44^ has recently emerged as a compelling alternative to convolutional U-Net architecture in diffusion models, harnessing the Transformer architecture’s ability to model long-range dependencies and to scale effectively in large-scale training. In this framework, images or other high-dimensional data are patchified into sequences and processed by Transformer blocks at each diffusion step, conditioned on timestep embeddings and optional label embeddings. This design allows DiT to capture both fine-grained local structure and global context, leading to improved generation quality and strong scalability. Recent studies have highlighted the effectiveness of DiT in high-resolution image synthesis, video generation, and multimodal applications such as text-to-image modelling, positioning it as a competitive backbone compared with U-Net–based designs.

### Model Pretraining: Implementation details

GenBrain is built on the DiT architecture. Unlike approaches that employ a pre-trained variational autoencoder to project brain images into a latent space, our model directly operates on registered brain voxels (Number of voxels = 228,453). A convolutional layer is used to patchify voxels into tokens, which are then passed into a 24-layer DiT backbone (depth = 24, hidden size = 1,024, patch size = 256, number of heads = 16). The model was trained for 200 epochs, corresponding to over 1.2 million iterations, with a learning rate of 1×10⁻⁴ using the AdamW optimizer^45^. Conditioning was provided via timestep embeddings together with age, sex, and modality index. Training was performed on 6 NVIDIA A100 80GB GPUs with an effective batch size of 192. The inference used DDIM approach with 50 denoising steps.

### Evaluation of the Pretrained GenBrain Model: Age-Sex Pattern Preservation

To evaluate how well GenBrain preserves age- and sex-related patterns, we compared the demographic correlation maps of synthetic images against a reference standard. Our method was as follows: First, we established a reference by selecting 18,345 subjects from the pre-training dataset with complete 34-modality coverage. For each modality, we computed the voxel-wise *t*-statistic maps between image intensity and age, and between image intensity and sex. This produced modality-specific reference spatial maps that characterize robust demographic patterns. Next, we used GenBrain to generate a full set of 34-modality images for randomly selected subjects (N = 100) from the hold-out dataset. We computed age and sex correlation maps from these synthetic images and measured their similarity to the reference maps using cosine similarity. To establish a confidence interval, we repeated this comparison five times using non-overlapping sets of real images (N = 100) from the hold-out dataset. The mean and variance of these cosine similarities define the expected range for real data.

### Evaluation of the Pretrained GenBrain Model: Detection of Training Data Replication

To determine whether GenBrain merely memorized and reproduced images from its UK Biobank pre-training dataset, we conducted an image-retrieval experiment. We first enumerated all 79 unique age–sex label combinations in the pre-training dataset and used them to condition GenBrain, generating a corresponding set of images across all 34 modalities via direct sampling. For each generated image, we extracted the brain voxels and computed its Pearson correlation with every real image of the same modality in the pre-training dataset. Any generated–real image pair with a correlation exceeding 0.95 was flagged as a potential instance of memorization.

### Fine-tuning for downstram tasks: Image enhancement

To demonstrate GenBrain’s adaptability for image enhancement, we fine-tuned it for denoising and motion removal on T1-weighted (T1w) and T2-FLAIR modalities.

To enable image-conditioned generation in the pretrained GenBrain, we replaced its original single-channel patch embedding layer with a two-channel layer. The corrupted image and the original input were concatenated along the channel dimension to form a two-channel input tensor. The model was then conditioned solely on the modality index, with age and sex information removed. GenBrain was fine-tuned for 50,000 steps on a simulated dataset of 2,000 paired samples (corrupted and clean), using a batch size of 64 and a learning rate of 1×10⁻⁴. For inference on each corrupted image, we sampled from the model five times and averaged the outputs to produce the final enhanced image.

Image corruption comprised motion artefacts and imaging noise. For motion artefacts, we employed a k-space–based model^46^ for simulation. In-acquisition motion was modelled as a sequence of rigid movements, each represented by a 3D affine transform (rotation and translation in the image domain). Rotation angles were constrained to [−30^∘^, 30^∘^] and translations to [−10,10] mm along the x, y, and z axes. Rotation and translation magnitudes were drawn from Poisson distributions (rotation: *λ* = 10; translation: *λ* = 5), assigned random signs, and truncated to the specified ranges. The number of motion events was sampled uniformly from (), and their occurrence times within k-space were also sampled uniformly. For imaging noise, we added Gaussian and Rician noise components at varying intensities, both of which are commonly observed in MR images^10^. Noise levels were controlled by the standard deviation *σ*, sampled independently from (0,5,10, …, 50}), and applied directly to the raw, unnormalized images.

We compared the fine-tuned GenBrain against several baselines: (1) GenBrain-st: A version of GenBrain trained from scratch on the same task without using the pre-trained weights. (2) Pix2Pix^21^: A leading generative adversarial network adapted for 3D medical imaging by replacing its 2D convolutional layers with 3D equivalents. It was trained under the same settings as the fine-tuned GenBrain. (3) BME-X^10^: A state-of-the-art foundation model for T1w MRI enhancement. We up-sampled corrupted images to a 1 mm isotropic resolution to match its input domain before processing. (4) SynthSR^22-24^: A tool that standardizes clinical brain scans into high-resolution, isotropic 1 mm T1w images and can enhance images with limited signal-to-noise ratios. Since SynthSR synthesizes the skull, we applied FreeSurfer’s SynthStrip^47^ for skull removal and SynthMorph^48^ for registration to the MNI152 2mm standard space (see Supplementary Note 2 for details).

Model performance was assessed using: (1) Image-level metrics: Peak signal-to-noise ratio (PSNR), root mean squared error (RMSE), 3D structural similarity index measure (3D-SSIM), and multi-scale SSIM (MS-SSIM). Before computation, all T1w and FLAIR images were normalized to the range [0,1]. PSNR and RMSE were calculated using brain voxels only, while 3D-SSIM and MS-SSIM were computed in the MNI152 2mm standard space. (2) Biological semantic-level metrics: Age-similarity and sex-similarity, defined as the cosine similarity between the age and sex correlation spatial maps of the enhanced images and the reference vectors, established during the pre-training model evaluation.

### Fine-tuning GenBrain for downstram tasks: Cross-modality synthesis

We demonstrate GenBrain’s adaptability by fine-tuning it for three distinct cross-modality synthesis tasks: (1) Structural Synthesis: between T1-weighted (T1w) and T2-FLAIR modalities. (2) Functional Synthesis: from resting-state fMRI (rs-fMRI) functional connectivity maps to task-based fMRI activations. Sources were 15 seed-based rs-fMRI functional connectivities (Rest-1 to Rest-15). Targets were shapes, faces task contrast maps (Task-1, Task-2). (3) Structure-Function Synthesis: from diffusion MRI scalar maps to functional connectivities. Sources comprised nine diffusion MRI scalar maps (DTI: FA, MD, MO, L1, L2, L3 and NODDI: OD, ICVF, ISOVF). Targets were functional connectivities for the Language Network (Rest-9) and the Visual Central Network (Rest-13). For all three tasks, each target were trained and evaluated in a separate experiment.

The fine-tuning procedure mirrored that used for image enhancement. To condition the model on multiple input images, we replaced the original single-channel patch embedding layer with a multi-channel layer. Age and sex conditions were removed, with only the target-modality index provided. For each experiment, we used 1,000 paired samples from the GenBrain pre-training dataset for training and 500 pairs from the held-out UK Biobank validation set for testing. Models were fine-tuned for 50,000 steps with a batch size of 32 and a learning rate of 1×10⁻⁴. During inference, we generated the final synthesized image by averaging five sampling outputs from GenBrain.

We compared the fine-tuned GenBrain against the following baselines: (1) TUMSyn^25^: A text-guided universal MRI synthesis model. We applied its provided checkpoint directly to the T1w/T2-FLAIR synthesis task without additional training. (2) U-Net: A 3D convolutional U-Net implemented via the MONAI framework^49^, with the number of input channels set to match the number of source modalities. (3) Pix2Pix^21^: A generative adversarial network adapted for 3D by replacing its 2D convolutional layers with 3D equivalents. Its input channels were also configured to match the number of source modalities. (4) GenBrain-st: An ablated version of our model trained from scratch on the synthesis tasks, without leveraging pre-trained weights. (5) SynthSR: the model was used only for FLAIR-to-T1w synthesis. U-Net, Pix2Pix, and GenBrain-st were all trained under the same settings as the fine-tuned GenBrain.

### Fine-tuning GenBrain for downstram tasks: Enhancing the reliability of brain-wide association study

Brain-wide association studies (BWAS) aim to identify voxels that show statistically significant differences between a group of healthy control subjects and a group of patients. To enhance the reliability of BWAS, particularly when dealing with small, single-site cohorts, we investigated whether GenBrain could be fine-tuned to generate realistic disease-specific brain images. The synthetic data were then used to perform BWAS, and the results were benchmarked against a large, multi-site reference cohort.

We conducted analyses across three distinct mental disorders and neuroimaging modalities to test the generalizability and adaptability of our approach: (1) Schizophrenia (SCZ): A multi-site dataset comprising 17 sites, using Voxel-Based Morphometry (VBM) as the target modality. (2) Major Depressive Disorder (MDD): A multi-site dataset comprising 24 sites, also using VBM. (3) Autism Spectrum Disorder (ASD): A multi-site dataset comprising 36 sites, using seed-based functional connectivity (FC) maps derived from the Tian subcortical atlas^26^ (16 regions). This modality is entirely distinct from those used during pre-training, providing a robust test of GenBrain’s adaptability to new image types. We present BWAS results from two of the 16 seed-based FC maps(right caudate nucleus and left anterior thalamus, Fig.5c). To improve interpretability, relative performance gains are reported in the Fig.5, whereas the corresponding raw performance metrics are provided in Supplementary Table 4.

To adapt GenBrain for BWAS, we introduced a disease-label embedder. For the ASD dataset, we further incorporated a new modality index to accommodate the functional connectivity maps. The model was conditioned on age, sex, and disease status. For each disorder, we selected representative sites from the corresponding multi-site dataset for fine-tuning. The model was trained on each single-site dataset for 10,000 steps with a batch size of 32 and a learning rate of 1×10⁻⁴.

Data from all remaining sites for a given disorder were pooled to form a large-scale reference cohort. This cohort was substantially larger than any single-site dataset and served as a gold standard for benchmarking reproducibility. After fine-tuning, GenBrain was used to generate synthetic samples conditioned on the metadata (age, sex, disease status). We generated a number of synthetic samples equivalent to the size of the reference cohort for a balanced comparison.

BWAS was performed separately on the real single-site data and the synthetic data generated to mimic that site. For VBM data, an average template was derived from the multi-site schizophrenia dataset with a threshold of 0.2. Association tests were performed within this common space. For all analyses, age and sex were included as confounding factors in the association tests for both the single-site and synthetic data. For the reference cohort, which combines multiple sites, age, sex, and site were regressed out in the association test to account for multi-site variability.

The performance was evaluated by comparing the Cohen’s *d* effect size statistical maps from the single-site (real or synthetic) data against the maps from the large reference cohort. Two primary metrics were used: (1) Pearson Correlation: The correlation between the vectorized BWAS maps from the site-level data and the reference cohort. (2) Dice Coefficient: A measure of voxel-wise overlap. Dice was calculated separately for voxels showing positive and negative effects. A weighted Dice score was defined as the micro-average of these two values. All Dice metrics were reported for the entire brain and for the top 20% of voxels ranked by their absolute Cohen’s *d* value.

### Fine-tuning GenBrain for downstram tasks: Data augmentation for machine learning disease diagnosis

To evaluate GenBrain’s adaptability and its utility in enhancing machine learning-based disease diagnosis, we fine-tuned the model on small, disease-specific cohorts. The objective was to test whether synthetic images generated by the fine-tuned model could improve diagnostic classification performance in both within-site and cross-site prediction scenarios.

We fine-tuned GenBrain on clinical data, conditioning the model on age, sex, and disease status. A disease-label embedder was introduced to represent diagnostic categories, where 0 denoted a healthy control and 1 denoted a diseased subject. The model was trained for 10,000 steps with a batch size of 32 and a learning rate of 1×10⁻⁴. We used T1-weighted (T1w) images as the target modality due to their widespread clinical use. Following training, synthetic T1w images were generated for subsequent analysis.

From each synthetic and real T1w image, we segmented 32 brain structures using WMH-SynthSeg^50^ and extracted their volumetric measures (Supplementary Table 5). These volumetric features served as quantitative inputs for a diagnostic classification model. We employed LightGBM, a gradient boosting framework based on tree-structured algorithms, to perform the classification. To systematically assess the impact of synthetic data on model performance, we varied the ratio of synthetic-to-real data in the training set from 0:1 (real data only) to 3:1, in increments of 0.1. For each ratio, the classification experiment was repeated 20 times with randomly selected combinations of LightGBM hyperparameters (e.g., number of estimators, maximum tree depth; see Supplementary Table 6).

We conducted experiments on two multi-site neuroimaging datasets. The schizophrenia (SZ) dataset is a multi-site dataset comprising data from 17 sites. We selected 7 sites that each contained more than 100 subjects with both cases and controls (site information see Supplementary Table 3). Within each site, data were split into training, validation, and test sets in a 3:1:1 ratio. For the Alzheimer’s disease (AD) datasets, we used T1w images from the ZIC dataset (N = 869), including Normal Control (NC), Mild Cognitive Impairment (MCI), and AD subjects. The data were split into training (N = 521), validation (N = 174), and test (N = 174) sets (3:1:1 ratio). For cross-site validation, we used the ADNI dataset (N = 2,632), partitioned into training (N = 1,562), validation (N = 554), and test (N = 516) sets using the same ratio.

For each disease-specific experiment, the optimal LightGBM classifier was selected based on the highest accuracy on the validation set. Model performance was then evaluated in two settings. For within-site prediction, performance was assessed on the held-out test set from the same site. For cross-site prediction, the generalizability was assessed by training on one site and evaluating on the full dataset of a different, unseen target site.

### Fine-tuning GenBrain for downstram tasks: Clinical Application

A critical test of a medical AI model’s utility is its robustness and generalizability across held-out datasets, particularly when the model is applied to patient populations and imaging protocols not seen during pre-training. This is especially relevant for GenBrain, as its pre-training stage utilized data from healthy participants. To evaluate its clinical applicability, we fine-tuned GenBrain on two distinct stroke-related prediction tasks and assessed whether synthetic data could enhance the performance of diagnostic classifiersFor NIH Stroke Scale (NIHSS) prediction in acute stroke, we collected 1,106 DTI–ADC scans from the SOOP dataset. All scans were registered to MNI152 2 mm standard space, and within-subject z-score normalization was applied. The dataset was split in a 3:1:1 ratio into a training set (N = 663), a validation set (N = 222), and a test set (N = 221). GenBrain was fine-tuned on the training set for 20,000 steps with a batch size of 32 and a learning rate of 1×10⁻⁴, using an extended modality index and disease labels.

For WAB Aphasia Quotient (WAB-AQ) prediction in chronic stroke, we used diffusion-weighted MRI from the ARC dataset (N = 213) and extracted fractional anisotropy (FA) maps. The dataset was split 3:1:1 into a training set (N = 129), a validation set (N = 41), and a test set (N = 43). GenBrain was fine-tuned on this data for 10,000 steps with a batch size of 8 and a learning rate of 1×10⁻⁴.

For both tasks, the synthetic-to-real data ratio was varied from 0:1 to 3:1. For each ratio, a 3D ResNet was trained for 50 epochs. This process was repeated five times with different random seeds, and the final performance was reported as the average across these runs. The 3D ResNet was trained with a learning rate of 5×10⁻⁵ and a batch size of 32 for the NIHSS task and 8 for the WAB-AQ task.

### Statistical analysis

For image enhancement and cross-modality synthesis tasks, image-level metrics, such as PSNR and 3D-SSIM, were computed, and the metric differences between GenBrain and other methods were assessed using paired two-sided t-tests. Biological semantic-level metrics, including age-similarity and sex-similarity, were evaluated using bootstrap t-tests (1,000 random resampling with replacement). Spatial correlations were calculated for each synthetic–real image pair, and the mean was used as the final metric, with differences tested using paired t-tests (two-sided). For BWAS, correlations and Dice coefficients were computed, and significance was determined using bootstrap t-tests (1,000 random resampling with replacement) within each cohort. In cross-site diagnosis, diagnosis performance across sites was evaluated using Accuracy and AUROC, with differences tested using bootstrap t-tests (1,000 random resampling with replacement). For clinical prediction tasks, including acute stroke severity and chronic aphasia quotient, diagnosis models were trained with five random seeds, and performance was measured using mean absolute error (MAE) and correlation; the mean across five runs was reported, and differences were assessed using two-sided t-tests. Error bars in figures represent 95% confidence intervals, calculated as 1.96 ×the standard error.

