## Supplementary for "GenBrain: A Generative Foundation Model of Multimodal Brain Imaging"

### Contents

**Supplementary Note 1 | Extending GenBrain to other non-imaging variables**

**Supplementary Note 2 | Image enhancement-SynthSR pipeline**

**Supplementary Note 3 | White matter hyperintensities analysis**

**Supplementary Note 4 | Fine-tuning GenBrain for image super-resolution**

**Supplementary Note 5 | Fine-tuning GenBrain for cross-modality synthesis (MNI152 1 mm standard space)**

**References**

**Supplementary Table 1 | Detailed information of 3D brain images across 34 modalities from the UK Biobank.** This table provides detailed information including **modality name, index, and description**. Abbreviations: **DTI** (Diffusion Tensor Imaging), **NODDI** (Neurite Orientation Dispersion and Density Imaging), **rs-fMRI** (Resting-state functional MRI), and **FC** (functional connectivity).

| Modality Name | Index | Description |
|---|---|---|
| DTI-FA | 0 | DTI-Fractional anisotropy |
| DTI-L1 | 1 | DTI-Axial diffusivity |
| DTI-L2 | 2 | DTI-Radial diffusivity |
| DTI-L3 | 3 | DTI-Second radial diffusivity |
| DTI-MD | 4 | DTI-Mean diffusivity |
| DTI-MO | 5 | DTI-Mode of anisotropy |
| NODDI-ICVF | 6 | NODDI-Intracellular volume fraction |
| NODDI-ISOVF | 7 | NODDI-Isotropic volume fraction |
| NODDI-OD | 8 | NODDI-Orientation dispersion |
| QSM | 9 | Quantitative susceptibility mapping from SWI |
| SWI | 10 | Susceptibility-weighted imaging |
| T1w | 11 | T1-weighted MRI |
| T1-Jac | 12 | Jacobian map of T1w nonlinear registration |
| T2-FLAIR | 13 | T2-Fluid attenuated inversion recovery |
| T2star | 14 | T2* from SWI |
| VBM | 15 | Grey matter volume from voxel-based morphometry |
| Task-1 | 16 | Task-based fMRI: Shapes contrast z-statistic maps |
| Task-2 | 17 | Task-based fMRI: Faces contrast z-statistic maps |
| Task-5 | 18 | Task-based fMRI: faces > shapes contrast z-statistic maps |
| Rest-1 | 19 | rs-fMRI seed-based FC: Visual Peripheral |
| Rest-2 | 20 | rs-fMRI seed-based FC: Cingulo-Opercular |
| Rest-3 | 21 | rs-fMRI seed-based FC: Default Network-B |
| Rest-4 | 22 | rs-fMRI seed-based FC: Somatomotor-B |
| Rest-5 | 23 | rs-fMRI seed-based FC: Auditory |
| Rest-6 | 24 | rs-fMRI seed-based FC: Premotor-Posterior Parietal Rostral |
| Rest-7 | 25 | rs-fMRI seed-based FC: Dorsal Attention-B |
| Rest-8 | 26 | rs-fMRI seed-based FC: Somatomotor-A |
| Rest-9 | 27 | rs-fMRI seed-based FC: Language |
| Rest-10 | 28 | rs-fMRI seed-based FC: Frontoparietal Network-B |
| Rest-11 | 29 | rs-fMRI seed-based FC: Frontoparietal Network-A |
| Rest-12 | 30 | rs-fMRI seed-based FC: Dorsal Attention-A |
| Rest-13 | 31 | rs-fMRI seed-based FC: Visual Central |
| Rest-14 | 32 | rs-fMRI seed-based FC: Salience / Parietal Memory Network |
| Rest-15 | 33 | rs-fMRI seed-based FC: Default Network-A |

**Supplementary Table 2 | Information on datasets used in this study**. Information about the number of sites (Site), subjects (Subject), scans (Scan), and image modality types (Modality) is provided. The UK Biobank dataset (used for pretraining and evaluation) contains 34 different image modalities; details are provided in Supplementary Table 1. For the ABIDE autism dataset, the Tian subcortical atlas was used to extract seed-based functional connectivity from the rs-fMRI data.

| Dataset | Site | Subject | Number of Scan | Modality |
|---|---|---|---|---|
| UK Biobank (pretraining) | 1 | 44,398 | 1,193,348 | 34 modalities |
| UK Biobank (evaluation) | 1 | 2,000 | 68,000 | 34 modalities |
| ZIC Alzheimer's disease | 1 | 1,138 | 2,271 | T1w, T2-FLAIR |
| ADNI | 1 | 768 | 2,632 | T1w |
| Schizophrenia | 17 | 2,958 | 5,780 | T1w, VBM |
| ABIDE I and II (ASD) | 36 | 1,778 | 1,778 | rs-fMRI |
| Major Depressive Disorder (MDD) | 24 | 2,831 | 2,831 | VBM |
| SOOP (acute stroke) | 1 | 1,106 | 1,106 | DTI-ADC |
| ARC (chronic stroke) | 1 | 213 | 213 | DTI-FA |

**Supplementary Table 3 | Details information of multisite datasets in our study.**

Information on the multisite datasets, including image modality (Modality), site name (Site), and number of subjects (Subjects), is provided.

| Dataset | Modality | Site Name (Subjects) |
|---|---|---|
| Schizophrenia | T1w | HCP-EP (93), ds004302(66), CLB (90), NUSDAST (250), chengdu (228), taiwan (254), ds000115 (40), ds000030 (171), NMorphCH (87), SH_JZ1 (298), COBRE (165), MCIC (203), zhengzhou (253), SH_JZ2 (324), SH_ECT (65), SH_drug1 (255), fBIRN (107) |
| | VBM | HCP-EP (93), ds004302 (64), CLB (90), NUSDAST (250), chengdu (103), taiwan (254), ds000115 (40), ds000030 (171), NMorphCH (87), SH_JZ1 (298), COBRE (165), MCIC (204), zhengzhou (253), SH_JZ2 (330), SH_ECT (65), SH_drug1 (257), fBIRN (107) |
| ABIDE I and II (ASD) | rs-fMRI | STANFORD (36), ABIDEII-GU_1 (99), ABIDEII-NYU_2 (27), ABIDEII-BNI_1 (54), ABIDEII-SDSU_1 (55), ABIDEII-OHSU_1 (86), YALE (48), CMU (5), ABIDEII-NYU_1 (72), NYU (171), LEUVEN_2 (32), OHSU (23), ABIDEII-EMC_1 (54), UM_2 (31), USM (61), ABIDEII-ETH_1 (34), ABIDEII-KKI_1 (198), SBL (26), ABIDEII-OILH_2 (58), UM_1 (82), ABIDEII-IU_1 (36), ABIDEII-KUL_3 (28), UCLA_1 (55), OLIN (25), PITT (45), TRINITY (44), MAX_MUN (42), ABIDEII-UCD_1 (13), LEUVEN_1 (29), UCLA_2 (20), ABIDEII-IP_1 (51), ABIDEII-TCD_1 (21), SDSU (33), KKI (39), ABIDEII-UCLA_1 (8), CALTECH (37) |
| Major Depressive Disorder | VBM | S1 (148), S2 (60), S3 (64), S5 (24), S6 (30), S7 (87), S8 (150), S9 (100), S10 (83), S11 (61), S12 (38), S13 (42), S14 (96), S15 (100), S16 (62), S17 (91), S18 (41), S19 (87), S20 (533), S21 (156), S22 (50), S23 (62), S24 (63), S25 (152) |

**Supplementary Table 4 | Raw BWAS performance metrics corresponding to Fig. 5.** Raw quantitative metrics underlying the relative improvements reported in Fig. 5, comparing real and synthetic data for brain-wide association studies (BWAS) across Schizophrenia (SCZ), Major Depressive Disorder (MDD), and Autism Spectrum Disorder (ASD). Metrics include Pearson correlation between voxel-wise Cohen's *d* maps, Dice coefficients computed on the full map and on positive ($d > 0$) and negative ($d < 0$) effects, as well as Dice coefficients restricted to the top 20% of absolute Cohen's *d* values. Relative improvements shown in Fig. 5 were computed from these raw values.

| Site | Disease | Data | Correlation | Dice | Dice-p | Dice-n | top-Dice | top-Dice-p | top-Dice-n |
|---|---|---|---|---|---|---|---|---|---|
| SH_JZ2 | SCZ | Real | 0.17 | 0.59 | 0.35 | 0.65 | 0.19 | 0.05 | 0.20 |
| SH_JZ2 | SCZ | Syn. | 0.29 | 0.69 | 0.37 | 0.77 | 0.29 | 0.08 | 0.30 |
| SH_JZ1 | SCZ | Real | 0.27 | 0.64 | 0.38 | 0.72 | 0.25 | 0.12 | 0.26 |
| SH_JZ1 | SCZ | Syn. | 0.30 | 0.68 | 0.38 | 0.76 | 0.27 | 0.18 | 0.28 |
| S20 | MDD | Real | 0.13 | 0.65 | 0.34 | 0.74 | 0.19 | 0.03 | 0.21 |
| S20 | MDD | Syn. | 0.18 | 0.67 | 0.37 | 0.76 | 0.19 | 0.06 | 0.21 |
| S9 | MDD | Real | 0.01 | 0.65 | 0.34 | 0.74 | 0.11 | 0.03 | 0.12 |
| S9 | MDD | Syn. | 0.01 | 0.67 | 0.37 | 0.76 | 0.16 | 0.04 | 0.17 |
| NYU | ASD | Real | 0.21 | 0.57 | 0.62 | 0.48 | 0.10 | 0.11 | 0.09 |
| NYU | ASD | Syn. | 0.27 | 0.60 | 0.67 | 0.48 | 0.31 | 0.35 | 0.09 |
| KKI_1 | ASD | Real | 0.16 | 0.68 | 0.77 | 0.32 | 0.23 | 0.24 | 0.01 |
| KKI_1 | ASD | Syn. | 0.22 | 0.71 | 0.81 | 0.34 | 0.25 | 0.25 | 0.03 |

**Supplementary Table 5 | Selected brain regions segmented using WMH-SynthSeg.** Region names and labels are listed, and their volumes were computed as features for the LightGBM classification task.

| Region | Label | Regions | Label |
|---|---|---|---|
| Left cerebral white matter | 2 | Left cerebral cortex | 3 |
| Left lateral ventricle | 4 | Left cerebellum white matter | 7 |
| Left cerebellum cortex | 8 | Left thalamus | 10 |
| Left caudate | 11 | Left putamen | 12 |
| Left pallidum | 13 | $3^{rd}$ ventricle | 14 |
| $4^{th}$ ventricle | 15 | Brainstem | 16 |
| Left hippocampus | 17 | Left amygdala | 18 |
| Extracerebral CSF | 24 | Left accumbens | 26 |
| Left ventral DC | 28 | Right white matter | 41 |
| Right cortex | 42 | Right lateral ventricle | 43 |
| Right cerebellum white matter | 46 | Right cerebellum cortex | 47 |
| Right thalamus | 49 | Right caudate | 50 |
| Right putamen | 51 | Right pallidum | 52 |
| Right hippocampus | 53 | Right amygdala | 54 |
| Right accumbens | 58 | Right ventral DC | 60 |
| WMH | 77 | Optic chiasm | 85 |

**Supplementary Table 6 | Hyperparameter settings of LightGBM**. For data augmentation in machine learning–based disease diagnosis, the classification experiment was repeated 20 times for each synthetic-to-real ratio using randomly selected combinations of LightGBM hyperparameters.

| Hyperparameter | Value | Description |
|---|---|---|
| n_estimators | {25,50,100,200,300} | Number of boosted trees to fit. |
| max_depth | {5, 10, 15, 20, 25, 30} | Maximum tree depth for base learners. |
| num_leaves | {5, 10, 15, 20, 25, 30} | Maximum number of leaves in one tree. |
| subsample | {0.60, 0.65, 0.70, …, 1.00} | Subsample ratio of the training instance. |
| learning_rate | {0.1, 0.05, 0.01, 0.001} | Learning rate. |
| colsample_bytree | {0.60, 0.65, 0.70, …, 1.00} | Subsample ratio of columns when constructing each tree. |
| boosting_type | {‘gbdt’} | Traditional gradient boosting decision tree. |

### Supplementary Note 1 | Extending GenBrain to other non-imaging variables

GenBrain was originally pretrained on the UK Biobank (UKB) dataset, using subject-specific biological variables—age and sex—as conditions for image generation. To examine whether GenBrain could also model images conditioned on other non-imaging phenotypic variables, we introduced fluid intelligence scores as an additional condition and fine-tuned the model on a subset of 1,000 subjects (26,939 images) from the UKB pretraining data. Fine-tuning was performed for 50,000 steps with a batch size of 64.

To evaluate the extent to which fluid intelligence–related patterns were preserved in the generated images, we followed the same procedure used to assess age- and sex-related biological patterns. Population-level pseudo–ground truth maps for fluid intelligence were constructed as voxel-wise t-statistic maps derived from a reference cohort (N = 18,345), with missing values imputed using the mean score (6.576).

Even when fine-tuned on only 1,000 subjects, GenBrain successfully captured fluid intelligence–associated spatial patterns in both structural MRI and task fMRI images. Quantitative evaluation results are provided in Supplementary Fig. 1.

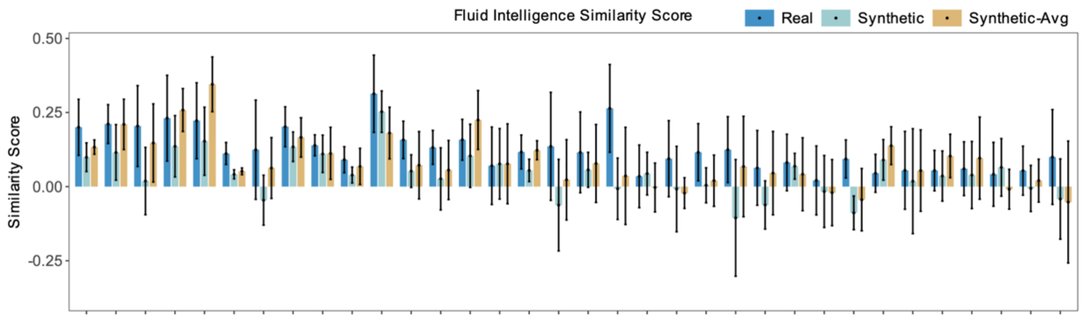


**Supplementary Fig. 1 | Preservation of fluid intelligence–related patterns in synthetic images.** GenBrain was fine-tuned on 1,000 subjects and extended to incorporate fluid intelligence scores as a new non-imaging conditional variable. The fluid intelligence–related pattern was evaluated on randomly sampled small cohorts (N = 100) of real data, synthetic data, and synthetic data averaged from five samples. The similarity score, defined as the cosine similarity between each small cohort's voxel-wise t-statistic map and that of the large reference cohort (N = 18,345), quantifies pattern preservation (higher scores indicate better preservation).

### Supplementary Note 2 | Image enhancement-SynthSR pipeline

SynthSR[1,2] is a tool that can standardize clinical brain scans into high-resolution, isotropic 1 mm T1w images and enhance low-field images with limited resolution and signal-to-noise ratios by using "--lowfield" option[3]. In the image enhancement task, we applied SynthSR to enhance the corrupted T1w image. The image processing pipeline consisted of three steps. First, SynthSR was used to super-resolve and synthesize 1 mm T1w images with the skull retained. The command used was:

```
mri_synthsr --i {corrupted_image} --o {image_with_skull} --threads {threads} --lowfield
```

Second, the skull was removed using FreeSurfer SynthStrip[4]. The command used was:

```
mri_synthstrip --i {image_with_skull} --o {brain_image} --threads {threads}
```

Finally, the skull-stripped images were registered to MNI152 2 mm standard space using FreeSurfer SynthMorph[5]. The command used was:

```
mri_synthmorph register -o {registered_image} -j {threads} -g {brain_image} {mni152_2mm}
```

In the above commands, all variables enclosed in curly brackets "{}", except for "`threads`", represent data paths. The "`registered_image`" denotes the final enhanced image. The number of processing threads in our experiment was set to 16. We also applied this pipeline by replacing "`corrupted_image`" with "`flair_image`" for the FLAIR-to-T1w cross-modality synthesis experiment.

**Supplementary Note 3 | White matter hyperintensities analysis**

In clinical practice, T2-FLAIR images are pivotal for detecting white matter hyperintensities (WMH), a neuroimaging biomarker widely associated with cognitive decline and an increased risk of Alzheimer's disease. Beyond achieving high performance on image-level and biological semantic-level metrics, high-fidelity synthetic T2-FLAIR images should also preserve clinically meaningful features such as WMH to ensure translational utility. To this end, we employed WMH-SynthSeg[6] to automatically segment and estimate the volume of WMH in the synthesized T2-FLAIR images (WMH label: 77).

For the cross-modality synthesis task, we performed WMH analysis on synthetic FLAIR images generated in the T1w-to-FLAIR task. WMH were first segmented, and their volumes were estimated in both real images and synthetic images generated by different methods. We then computed the correlation of WMH volumes between paired real and synthetic images to evaluate how well the synthetic images preserved WMH features.

In the inner-dataset evaluation, GenBrain-ft achieved the highest correlation score (0.928) on the UKB dataset (N = 500). In the external evaluation on the ZIC dataset (N = 200), TUMSyn and GenBrain-ft obtained similar best correlation scores (TUMSyn: 0.922; GenBrain-ft: 0.911). The strong generalization performance of TUMSyn can be attributed to its multi-dataset training strategy, whereas GenBrain-ft benefited from generative pretraining on the UKB dataset. Detailed results are provided in Supplementary Fig. 2.

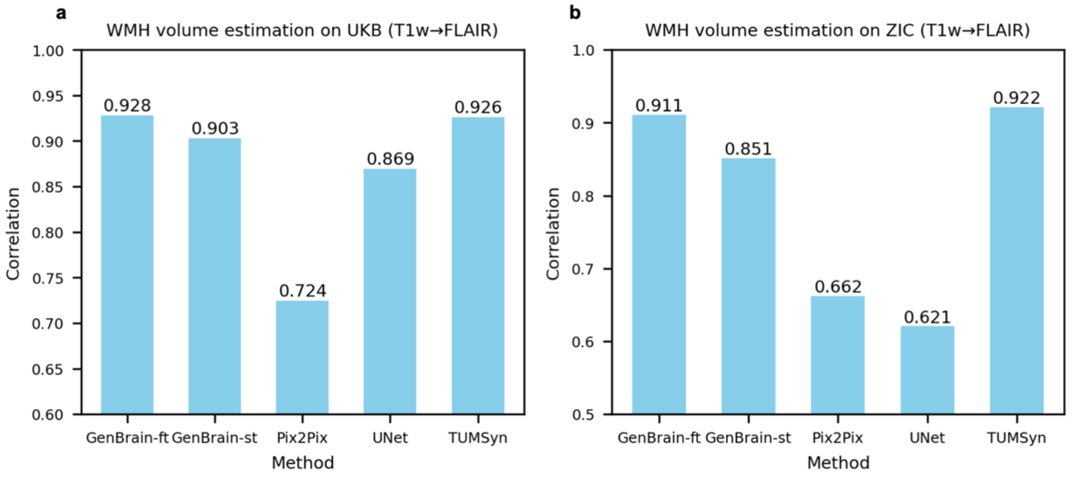


**Supplementary Fig. 2 | Correlation of WMH volumes between paired real and synthetic images.** WMH analysis was performed on synthetic FLAIR images in the T1w-to-FLAIR experiments. Internal dataset evaluation was conducted on the UKB dataset, and external evaluation on the ZIC dataset. High correlation indicates that the synthetic images reliably reproduce the WMH volume patterns existed in real T1w images.

**Supplementary Note 4 | Fine-tuning GenBrain for image super-resolution**

GenBrain was originally pretrained and analyzed in the MNI152 2 mm standard space (voxel size: $2 \times 2 \times 2\ mm^3$ ). To evaluate its adaptability to higher-resolution images, we fine-tuned the model to perform super-resolution from MNI152 2 mm to MNI152 1 mm standard space.

To adapt GenBrain for super-resolution, low-resolution (2 mm) T1-weighted images were first upsampled to 1 mm resolution using nearest-neighbor interpolation. Because directly applying the model's patch embedding to high-resolution images would incur a quadratic increase in computational cost due to the self-attention mechanism, we divided the interpolated images into eight equal-sized parts. Each part contained 228,453 voxels, the same number as in the corresponding low-resolution input, and was assigned a unique part index. The original single-channel patch embedding layer was replaced with a two-channel embedding layer, and a new image part–index embedder was introduced. This fine-tuned model is denoted as GenBrain-SR.

During fine-tuning, each interpolated part was concatenated with its corresponding noised high-resolution image part along the channel dimension to form a two-channel input tensor. GenBrain-SR was conditioned on this input tensor, the modality, and the part indices; age and sex embeddings were omitted. Fine-tuning was performed for 50,000 steps with a batch size of 64, using 1,000 paired T1w images (at both MNI152 1 mm and 2 mm resolutions) from the UK Biobank pretraining dataset.

During inference, GenBrain-SR generated high-resolution outputs part-by-part. These parts were subsequently combined into a complete high-resolution image by averaging the values of overlapping voxels. We found that GenBrain-SR could not only super-resolve real images but also further enhance synthetic images, such as the 2 mm T1w images initially generated by GenBrain-ft. Representative examples are provided in Supplementary Fig. 3.

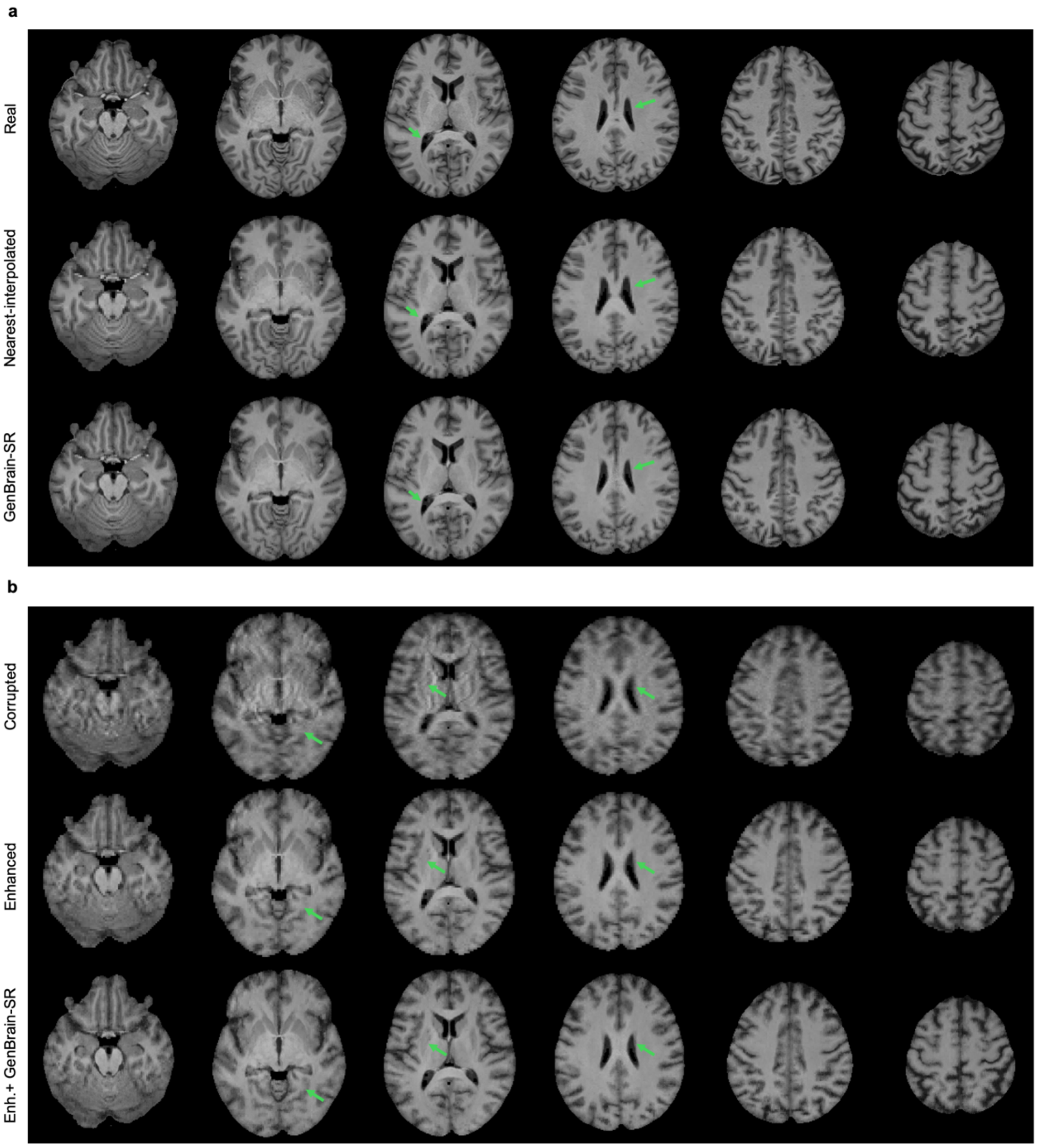


**Supplementary Fig. 3 | Fine-tuning GenBrain for image super-resolution (MNI152 2 mm → 1 mm). a**, Examples of high-resolution T1w images (1 mm), nearest-neighbor–interpolated T1w images (2mm → 1mm), and GenBrain-SR processed T1w images. **b,** Examples of corrupted T1w images (2 mm), GenBrain-ft–enhanced T1w images (2 mm), and the enhanced T1w images further super-resolved by GenBrain-SR (1 mm).

**Supplementary Note 5 | Fine-tuning GenBrain for cross-modality synthesis (MNI152 1 mm standard space).**

To further evaluate GenBrain's adaptability to higher-resolution images, we conducted T1w to T2-FLAIR and T2-FLAIR to T1w cross-modality synthesis tasks with images registered in MNI152 1mm standard space. Similar to the image super-resolution task, each source modality image was divided into eight parts of equal size, and GenBrain adopted the same architectural modifications, including a two-channel patch embedding layer and an image part–index embedder. GenBrain was conditioned on the source image, target modality and part indices, while age and sex embeddings were removed. GenBrain was fine-tuned for 50,000 steps with a batch size of 64 using 1,000 paired images (source–target modality pairs) from the UK Biobank pretraining dataset. At inference, the model translated the source images part-wisely, and then combined image parts into the target modality images (overlapped voxels using the average value). Cross-modality synthesis results see Supplementary Fig. 4.

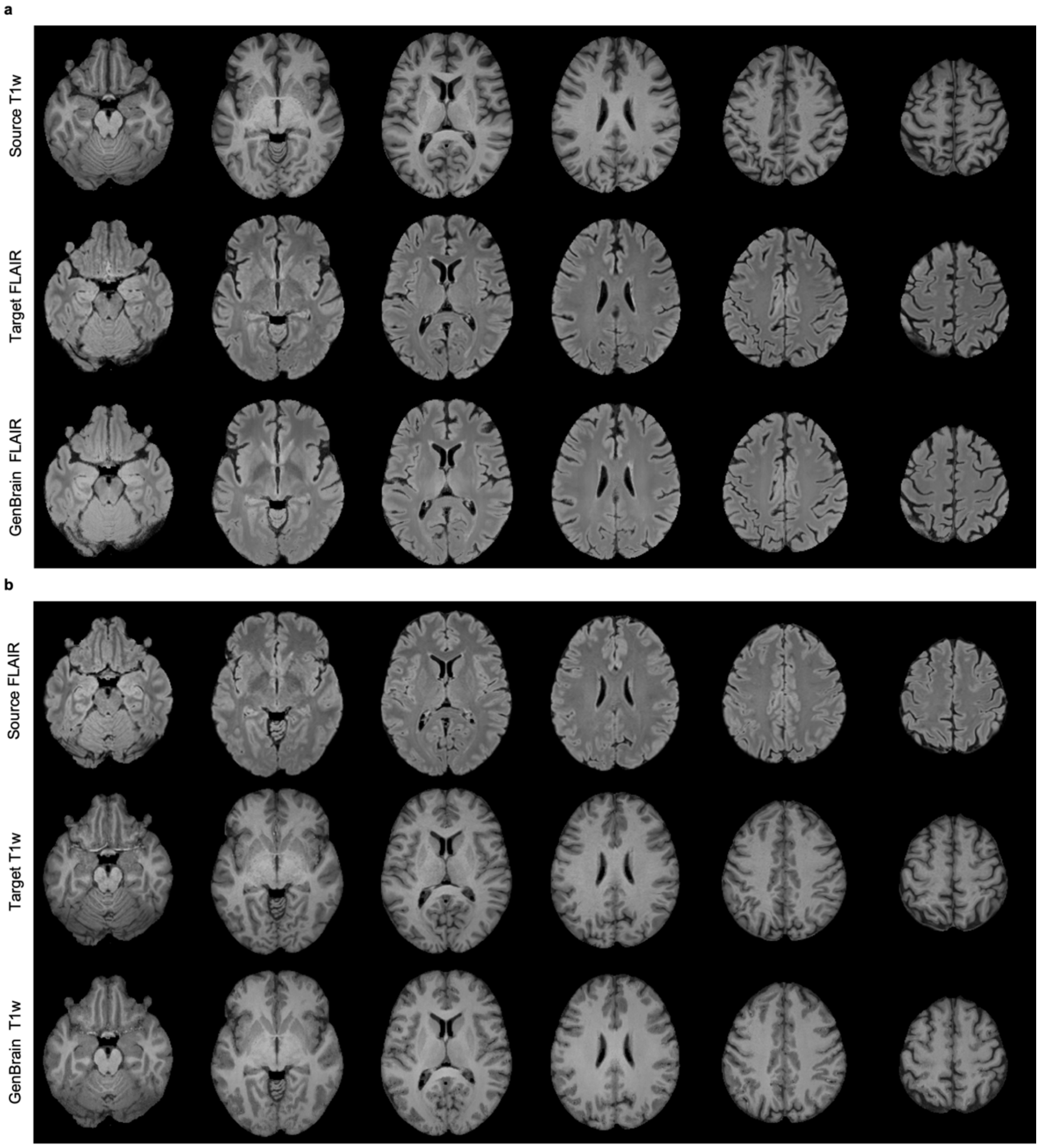


**Supplementary Fig. 4 | Fine-tuning GenBrain for cross-modality synthesis (MNI152 1 mm). a**, T1w to T2-FLAIR synthesis. Examples of source T1w images, target T2-FLAIR images, and GenBrain synthesized T2-FLAIR images are shown. **b**, T2-FLAIR to T1w synthesis. Examples of source T2-FLAIR images, target T1w images, and GenBrain synthesized T1w images are shown.
